# Declining but increasingly concentrated HIV stigma in rural Uganda: population-based cohort study, 2014-2024

**DOI:** 10.64898/2026.05.08.26352137

**Authors:** Alexander C. Tsai, Charles Baguma, Phionah Ahereza, Scholastic Ashaba, Patience Ayebare, David R. Bangsberg, Alison B. Comfort, Patrick Gumisiriza, Mercy Juliet, Justus Kananura, Allen Kiconco, Viola Kyokunda, Patrick Lukwago, Rumbidzai S. Mushavi, Elizabeth B. Namara, Jessica M. Perkins, Justin M. Rasmussen, Emily N. Satinsky, Mark J. Siedner, Benjamin Martin Tweheyo, Bernard Kakuhikire

**Affiliations:** Center for Global Health, Massachusetts General Hospital, Boston, Massachusetts, United States; Harvard Medical School, Boston, Massachusetts, United States; Department of Epidemiology, Harvard T.H. Chan School of Public Health, Boston, Massachusetts, United States; Mbarara University of Science and Technology, Mbarara, Uganda; Faculty of Health Sciences, Simon Fraser University, Burnaby, British Columbia, Canada; Department of Obstetrics and Gynecology, University of California at San Francisco, San Francisco, California, United States; Department of Obstetrics and Gynecology, Massachusetts General Hospital, Boston, Massachusetts, United States; Peabody College, Department of Human and Organizational Development, Vanderbilt University, Nashville, Tennessee, United States; Department of Psychology and Neuroscience, Duke University, Durham, North Carolina, United States; Department of Psychology; Dana and David Dornsife College of Letters, Arts, and Sciences; University of Southern California, Los Angeles, California, United States; Medical Practice Evaluation Center, Massachusetts General Hospital, Boston, Massachusetts, United States; Division of Infectious Diseases, Department of Medicine, Massachusetts General Hospital, Boston, Massachusetts, United States

**Keywords:** discrimination, eastern Africa, HIV, stigma, sub-Saharan Africa, Uganda

## Abstract

**Background:** HIV-related stigma remains a primary barrier to the elimination of the HIV epidemic worldwide. No studies have examined long-term changes in the distribution of stigmatizing attitudes within populations.

**Methods:** We conducted a whole-population, open cohort study of adults in 8 villages in rural southwestern Uganda, with 5 biennial surveys spanning 2014-2024 (N=1,776 at baseline; 869 participated in all waves). We measured individual negative attitudes toward people with HIV (“public stigma”) and perceptions of negative attitudes among others (“perceived stigma”) using parallel 15-item scales. We estimated mean stigma scores, computed inequality measures at each wave, and decomposed inequality by sociodemographic characteristics. Leveraging the cohort design, we estimated intraclass correlation coefficients and rank-order stability over time.

**Results:** Both public and perceived stigma declined substantially from baseline to endline and became concentrated in an increasingly smaller subgroup of the population. Theil decomposition failed to identify any stratifying variables that explained more than 3% of this variation: nearly all the inequality in HIV stigma occurred within population subgroups rather than between them. In longitudinal analyses, public stigma showed trait-like properties (intraclass correlation coefficient=0.35; 95% CI, 0.31-0.38) and meaningful rank-order stability (baseline-to-endline r=0.41). Perceived stigma showed no rank-order stability, no appreciable between-person variance, and universal convergence to low levels regardless of baseline.

**Conclusions:** Both public and perceived HIV stigma declined substantially in this rural Ugandan population, but remaining public stigma has become concentrated within a persistent minority. Sociodemographic profiling to target individuals who carry persistently negative attitudes toward people with HIV is unlikely to succeed.

---

HIV-related stigma has long been recognized as a fundamental barrier to eliminating the HIV epidemic (1–3). Surveys based on diverse sampling designs and in diverse settings have documented persistently negative attitudes toward people with HIV (4–11). This stigma is a problem for public health because it discourages individuals from seeking HIV testing and—among those who test positive—delays linkage to care, erodes the social support networks that facilitate health-seeking behavior, worsens emotional wellbeing, undermines adherence to antiretroviral therapy, and reduces the probability that viral suppression will be achieved (11–20).

Over the past several decades, substantial progress has been made in understanding the determinants and consequences of HIV stigma as well as how to effectively respond to it. Various approaches—including information provision, contact-based strategies, and counseling—can reduce negative attitudes toward people with HIV, though effect sizes are often modest and long-term durability is uncertain (3, 21–23). In many settings, negative attitudes toward people HIV have attenuated as HIV testing and treatment have expanded at scale (9, 24–26), but the findings from population trials have been mixed in this regard (27, 28).

Despite this progress, important gaps remain in our understanding of how HIV stigma has declined and the implications for further intervention. Most evidence on long-term stigma trends comes from repeated cross-sectional surveys, generally conducted over a several year period (9, 11, 29–32). There are important exceptions, including the Zambia/South Africa HPTN 071 (PopART) trial (27) and Maughan-Brown’s (33) analysis of data from the Cape Area Panel Survey, both of which followed the same set of participants over three years of follow up. Studies focused on people with HIV (who are typically engaged in care) offer opportunities for longer follow-up but are limited to eliciting the perspectives of people with HIV, with no ability to extrapolate to broader population-level trends (34–38).

Interpretation of the findings from serial cross-sectional studies is subject to two fundamental limitations. First, they cannot distinguish changes in HIV stigma from compositional shifts in the population: observed declines in mean stigma scores could reflect changes in negative attitudes toward people with HIV among community members, or the observed declines could simply differential attrition and population replacement. Second, serial cross-sectional studies cannot characterize heterogeneity in individual trajectories of stigma. Namely: when the mean level of HIV stigma declines in the population, are negative attitudes toward people with HIV proportionally reduced among everyone in the population, or are stigmatizing attitudes reduced substantially in a subset of the population but persist in another subset of the population? This distinction is important for public health because it carries direct implications for intervention design: uniform change suggests that population-level approaches are reaching everyone equally, while heterogeneous change implies that complementary targeted interventions may be needed for those who do not respond.

To address these gaps in the literature, we analyzed data from a whole-population cohort study in Mbarara, Uganda spanning a decade (2014–2024). We examined trends not only in mean HIV stigma scores but also in how stigma is distributed within the population, using well-established metrics of inequality from development economics, with the aim of determining the extent to which stigma is increasingly concentrated among identifiable sociodemographic subgroups. Our analysis leverages the cohort design, which enabled analyses not possible with a serial cross-sectional design.

## METHODS

### Study setting

This study was conducted in Nyakabare Parish, a rural area of 8 villages in Mbarara District, southwestern Uganda. The parish is located in a rural area approximately 20 kilometers from Mbarara City, which had a population of approximately 250,000 according to the Uganda Bureau of Statistics (39) census. The local economy is largely based on subsistence agriculture, animal rearing, and small-scale enterprise and trading; both food and water insecurity are common challenges in the region (40, 41). Uganda has experienced a generalized HIV epidemic: the prevalence of HIV in the Mbarara region was estimated at approximately 8% at the time our study was initiated, slightly higher than the national prevalence of 6% (42). In 2004 Uganda became the first country to provide antiretroviral therapy, supported by the U.S. President’s Emergency Plan for AIDS Relief, with progressive expansion of treatment eligibility culminating in the adoption of “treat all” guidelines in 2016 (43, 44).

### Study design and participants

We initiated a whole-population open cohort study of adults residing in the 8 study villages in 2014 (45). Several months before survey administration, study staff held “community sensitization” meetings throughout the parish (46). Then we enumerated all adults (age 18 years or older, or emancipated minors aged 16-18 years) residing within the parish who considered the parish their home. Of the 1,942 adults eligible for participation, 1,776 (91%) agreed to participate in the baseline survey (45).

Study interviews were done in-person. Survey questions were written in English, translated into Runyankore (the local language) and piloted following an iterative process, then back-translated to English to verify fidelity to the intended meaning. Survey data were captured electronically using the Computer Assisted Survey Information Collection Builder(TM) software program (West Portal Software Corporation, San Francisco, Calif., USA).

We conducted follow-up surveys biennially after baseline. At each follow-up wave, we attempted to re-interview all baseline participants and, following the open cohort design, also enrolled new residents who had moved into the study catchment area (**Appendix Figure S1**). Sample sizes at each survey wave are shown in **Appendix Table 1**. Data collection was temporarily halted on March 20, 2020, during the third survey wave, due to country-wide restrictions on internal movement implemented to suppress the spread of COVID-19. Data collection resumed April 21, with study interviews conducted by mobile phone. In-person interviews resumed on July 26, 2022, near the end of the fourth survey wave.

**Table 1.**
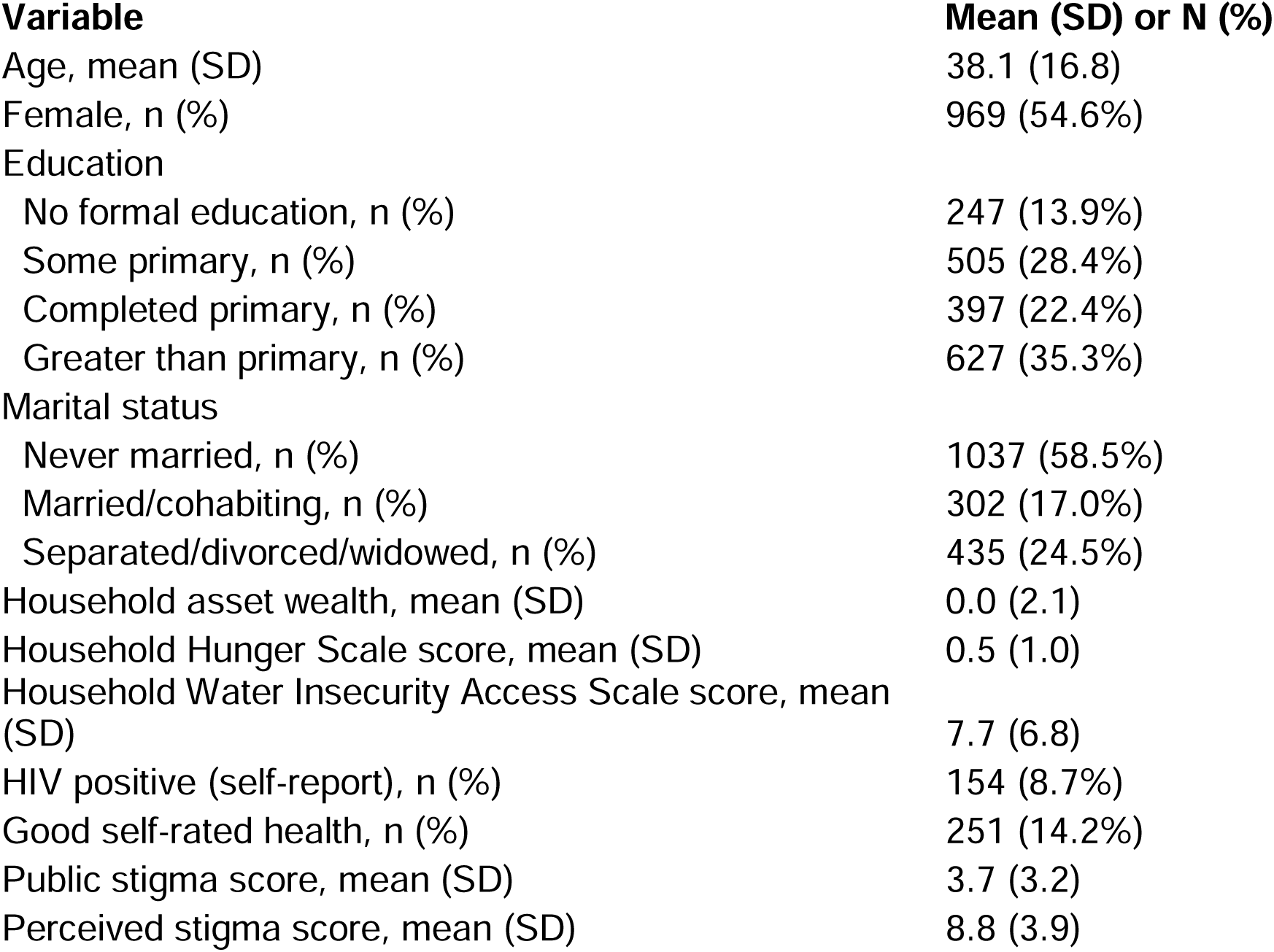
Characteristics of the study population, at baseline.

| **Variable** | **Mean (SD) or N (%)** |
| --- | --- |
| Age, mean (SD) | 38.1 (16.8) |
| Female, n (%) | 969 (54.6%) |
| Education |  |
| No formal education, n (%) | 247 (13.9%) |
| Some primary, n (%) | 505 (28.4%) |
| Completed primary, n (%) | 397 (22.4%) |
| Greater than primary, n (%) | 627 (35.3%) |
| Marital status |  |
| Never married, n (%) | 1037 (58.5%) |
| Married/cohabiting, n (%) | 302 (17.0%) |
| Separated/divorced/widowed, n (%) | 435 (24.5%) |
| Household asset wealth, mean (SD) | 0.0 (2.1) |
| Household Hunger Scale score, mean (SD) | 0.5 (1.0) |
| Household Water Insecurity Access Scale score, mean (SD) | 7.7 (6.8) |
| HIV positive (self-report), n (%) | 154 (8.7%) |
| Good self-rated health, n (%) | 251 (14.2%) |
| Public stigma score, mean (SD) | 3.7 (3.2) |
| Perceived stigma score, mean (SD) | 8.8 (3.9) |

Among the 1,776 participants interviewed at baseline, 1,052 (59%) were successfully reinterviewed at final follow-up; 869 (49%) participants were successfully interviewed at all 5 survey waves.

### Measurement of HIV stigma

HIV stigma was assessed using a 15-item scale developed for use in Mbarara (45). The scale captures three domains of negative attitudes toward people with HIV, which for convenience we refer to as “public stigma” following Corrigan and Watson (47). The first two domains—fear of acquisition through everyday contact and attributions of blame, judgment, and moral responsibility—are well known drivers of public stigma, understood through Pryor et al.’s (48) “instrumental” vs. “symbolic” typology. The third domain, representing concerns about the ability of people with HIV to make reciprocal contributions to local networks of mutual aid, was based on qualitative research conducted in Mbarara and elsewhere in eastern and southern Africa (45, 49, 50).

For each item, respondents were asked about both their own attitudes (public stigma) and their perceptions of prevailing negative attitudes in their communities (which for convenience we refer to as “perceived stigma”). For the public stigma items, study participants were asked to indicate agreement or disagreement with the statement. The public stigma index was constructed as the sum of item responses, with items appropriately recoded so that a 1 reflects a stigmatizing response, yielding a score ranging from 0 to 15. For the perceived stigma items, study participants were asked about their perceptions of the beliefs of others, with response options scored on a four-point Likert-type scale ranging from “very few, or no one” to “all or almost all.” Responses to the perceived stigma items were dichotomized such that 1 indicated the participant’s belief that “most people” or “everyone” in the community held the stigmatizing attitude. The perceived stigma index was similarly constructed as the sum of dichotomized items, yielding a score ranging from 0 to 15. All items permitted refusals and “don’t know” responses, which were minimal.

### Other covariates

Sociodemographic characteristics assessed at each wave included age (continuous), sex (female vs. male), educational attainment (no formal education, primary, secondary, or tertiary), and marital status. Household asset wealth was measured using a household asset index derived from principal components analysis applied to household possessions and dwelling characteristics (51, 52). Food security was assessed using the Household Hunger Scale (53), which is a 3-item subset of the 9-item Household Food Insecurity Access Scale (54, 55). Water insecurity was assessed using the 8-item Household Water Insecurity Access Scale (41, 56). Self-rated overall health was measured with a single item with responses scored on a four-point Likert-type scale ranging from “very bad” to “very good” (57). HIV serostatus was elicited by self-report (positive, negative, don’t know).

### Statistical analysis

We first conducted descriptive analyses treating the data as serial cross-sections. We calculated mean stigma scores and standard deviations at each wave for both public and perceived stigma. To quantify the statistical significance of any observed change over the study period, we fit a linear mixed effects regression model specifying HIV stigma as the dependent variable and survey wave as a covariate, interpreting the estimated coefficient on survey wave as the extent of change in HIV stigma per survey period.

To characterize the distributional properties of HIV stigma, we applied inequality metrics originally used in development economics (58, 59). We calculated the Gini coefficient, which ranges from 0 (perfect equality) to 1 (perfect inequality), and the Theil index, an entropy-based measure that is decomposable into between-group and within-group components. The Gini is more sensitive to changes around the middle of the distribution, while the Theil is more sensitive to changes in upper-tail concentration (60). For a more accessible metric, we also computed top decile and top quintile shares (61, 62), defined as the percentage of total stigma “burden” in the population held by the top 10% and 20% of individuals, respectively.

While these complementary measures were originally developed to measure income and wealth inequality, they can be meaningfully applied to any quantifiable health-related attribute distributed within a population, including height (63) and life expectancy (64). Their interpretation as measures of concentration (or dispersion) of income and wealth can be applied to characterizing the extent to which HIV stigma is distributed broadly within the population or highly concentrated within a minority. Our interpretation of these statistics differs from an analysis of income and wealth, however, in that high levels of inequality in income and wealth are normatively undesirable. Rising “inequality” of stigma observed during a period of relative decline could potentially represent a positive epidemiological transition, e.g., most community members have adopted less stigmatizing attitudes but the attitudes of a minority subgroup have remained refractory to change. This distributional perspective potentially complements analyses focused solely on measures of central tendency and dispersion by identifying the individuals with sustained stigmatizing attitudes even as population levels decline.

Next, we employed Theil decomposition (65) to assess the extent to which the observed concentration of HIV stigma reflects differences between population subgroups or variation among individuals within subgroups. The Theil index is uniquely suited to answering this question because, unlike the Gini coefficient, it can be exactly partitioned into between-group vs. within-group components (66). We conducted Theil decomposition separately for seven stratifying variables: age, sex, educational attainment, marital status, household asset wealth, food insecurity, water insecurity, self-rated health, and HIV status.

We then turned to an analysis of data restricted to the subset of 869 individuals who participated in all 5 survey waves. We estimated unconditional linear mixed-effects regression models (i.e., with only a random intercept for individuals), partitioning total variance in HIV stigma into between-person variance and within-person variance. The intraclass correlation coefficient (ICC) indicates the proportion of variance attributable to stable individual differences: an ICC near 1 indicates that stigma is trait-like, with individuals maintaining stable relative attitudes over time, while an ICC near 0 indicates that stigma is state-like, with attitudes fluctuating over time independent of any stable disposition. To avoid conflating within-person changes in stigma with population-level secular trends, we also estimated ICC by including survey wave as a fixed effect in the multilevel regression models. To assess rank-order stability, we also calculated Pearson correlations of stigma scores across all pairs of waves. High correlations indicate that study participants who harbored high levels of stigma at one time point tended to harbor high levels of stigma at subsequent time points; low or negative correlations indicate that relative rankings are not preserved.

### Sensitivity analyses

We conducted four sensitivity analyses to probe the robustness of our findings to potential confounding by variables omitted from consideration.

First, it is possible that the observed trends in stigma were driven by unobserved social desirability bias. Omission of this variable from the regression model could result in study participants reporting increasingly reduced stigma even if they continued to maintain persisting negative attitudes toward people with HIV. To assess for this possibility, we estimated e-values (67) for the regression models estimating the change in stigma per survey wave.

Second, potential concern with serial cross-sectional designs is that observed trends in stigma could reflect compositional shifts resulting from different people being sampled at different time points. For example, individuals with more stigmatizing attitudes and more life stability may be differentially more likely to migrate out of the study catchment area, causing us to underestimate the extent of concentration over time. We compared the baseline characteristics of the 869 retained study participants against the 907 study participants who were not retained. As an additional sensitivity check (and to ensure that any observed trends could not be driven by compositional shifts), we replicated the inequality analyses restricted to the balanced panel of 869 individuals who participated in all five survey waves.

Relatedly, a potential problem with the ICC and rank-order stability analyses is that if the balanced panel is comprised of individuals with less stigmatizing attitudes and/or less life instability (i.e., people with more stigma and more life instability are differentially lost to follow-up), the ICC estimates could be upwardly biased. To address this potential selection bias due to differential attrition, in a fourth sensitivity analysis we used inverse probability of censoring (IPC) weighting.

All analyses were conducted using R version 4.3. For descriptive statistics, including the inequality measures and correlation coefficients, we computed 95% confidence intervals using nonparametric cluster bootstrap with 1,000 replications, resampling villages with replacement to account for clustering of participants within villages (68). In regression models, we accounted for clustering of participants within villages with cluster robust (CR2) standard errors using small sample adjustments implemented in the clubSandwich package in R (69, 70).

### Ethics approval

All research assistants underwent intensive training in fieldwork procedures and survey administration for collecting sensitive information, including guidance on how to pause the interview if another person came within earshot of the interview. They also received two supplementary trainings—one from The AIDS Support Organization and one from a Ugandan counseling psychologist—focused on responding to sensitive disclosures from study participants. Each eligible individual was approached in the field, usually at their home or workplace, by a research assistant fluent in the local language (Runyankore). The research assistant briefly presented the study as a community survey about the social lives and health of residents of Nyakabare Parish (i.e., not as a study specifically about attitudes toward people with HIV) and invited them to take part. For individuals who indicated possible interest, the study was then explained in full, and written informed consent was obtained. Participants who were unable to write were allowed to provide consent via thumbprint.

We sought input on the study design from a community advisory board consisting of eight community leaders (four women and four men), including the district community development officer and an HIV-positive volunteer counselor from the Mbarara Regional Referral Hospital Immune Suppression Syndrome Clinic. In addition, we held a series of community sensitization meetings in each study village, during which we described the study design in detail, responded to questions, and invited feedback (46).

All study participants provided written informed consent. Consistent with local custom, they received a bar of soap or a kilogram of sugar as a study participation incentive. Ethical approval for all study procedures was granted by the Mbarara University of Science and Technology and the Partners Human Research Committee. During the COVID-19 pandemic, these review boards authorized a modification from written informed consent to verbal consent procedures. In accordance with national guidelines, we also obtained permission to conduct the study from the Uganda National Council of Science and Technology and from the Research Secretariat in the Office of the President.

## RESULTS

### Characteristics of the sample

Characteristics of the study population at baseline are shown in **Table 1**. The population was relatively young, with a mean age of 38 years, and had a higher percentage of women than men. Approximately one-third had completed a primary education or more. Self-reported HIV prevalence was 8.7%. Characteristics of the study population over the course of the decade are shown in **Appendix Table S1**. A flow diagram depicting the open cohort design is shown in **Appendix Figure S1**.

### Changes in HIV stigma and its distribution over time

At baseline, the mean public stigma score was 3.7 (standard deviation [SD], 3.2), while the mean perceived stigma score was 8.8 (SD, 3.9). Both the mean and variance of public stigma and perceived stigma declined substantially over the study period (**Figure 1A**). Public stigma declined 61% in relative terms, or 0.7 standard deviation units; in the mixed-effects linear regression model, the estimated coefficient was b=-0.59 per wave (95% confidence interval [CI], –0.67 to –0.52). Perceived stigma declined 73% in relative terms, or 1.6 standard deviation units; the estimated regression coefficient was b=-1.70 per wave (95% CI, –1.78 to –1.62).

**Figure 1.**
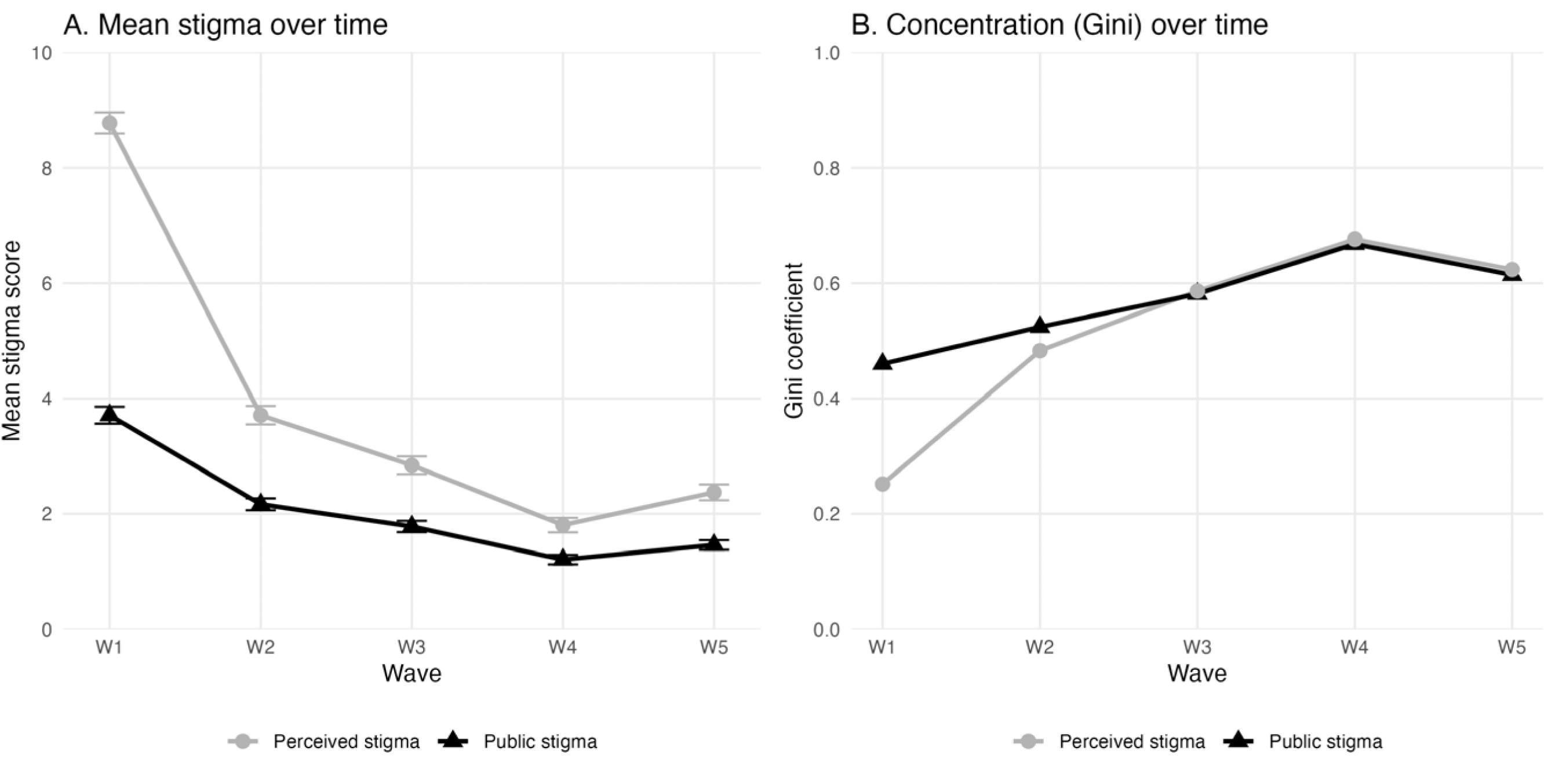
Trends in mean stigma (Panel A) and concentration (Panel B), 2014-24.

While average levels of stigma declined overall, its distribution in the population changed substantially (**Figure 1B** and **Appendix Figure S2**). The Gini, Theil, and top decile/quintile shares all increased (**Table 2**). Perceived stigma declined more steeply, and more unequally, than public stigma. The greater rates of change in the Theil index confirm that stigma was becoming more concentrated in the upper tail: a subgroup of the population retained high levels of stigma while the remainder the population converged toward zero.

**Table 2.** Trends in mean stigma and concentration, 2014-24.

| Wave | N | Mean (SD) | %<br>None | Gini* | Top Decile<br>Share* | Top Quintile<br>Share* | Theil* |
| --- | --- | --- | --- | --- | --- | --- | --- |
| *Panel A. Public stigma* |  |  |  |  |  |  |  |
| W1 | 1741 | 3.74 (3.17) | 11.4 | 0.458 [0.441, 0.471] | 28.4 [26.7, 30.2] | 47.7 [45.3, 49.4] | 0.363 [0.334, 0.388] |
| W2 | 1335 | 2.11 (2.11) | 27.2 | 0.534 [0.515, 0.553] | 31.9 [30.4, 33.4] | 52.6 [50.5, 54.8] | 0.526 [0.486, 0.567] |
| W3 | 1219 | 1.74 (2.00) | 34.4 | 0.592 [0.571, 0.615] | 36.2 [34.5, 38.1] | 58.7 [56.9, 61.4] | 0.654 [0.606, 0.715] |
| W4 | 1132 | 1.21 (1.67) | 49.2 | 0.670 [0.653, 0.684] | 42.6 [40.7, 44.1] | 65.8 [64.1, 67.2] | 0.880 [0.826, 0.926] |
| W5 | 1038 | 1.37 (1.72) | 44.1 | 0.636 [0.613, 0.660] | 38.3 [36.5, 40.6] | 62.2 [59.8, 65.3] | 0.778 [0.718, 0.848] |
| *Panel B. Perceived stigma* |  |  |  |  |  |  |  |
| W1 | 1741 | 8.80 (3.91) | 4.9 | 0.250 [0.230, 0.267] | 16.6 [15.9, 16.9] | 31.1 [30.1, 32.0] | 0.129 [0.110, 0.148] |
| W2 | 1335 | 3.56 (3.18) | 18.7 | 0.489 [0.474, 0.503] | 28.4 [27.8, 29.2] | 48.2 [47.1, 49.4] | 0.428 [0.401, 0.454] |
| W3 | 1219 | 2.78 (3.18) | 31.7 | 0.596 [0.582, 0.610] | 35.6 [33.9, 37.6] | 58.1 [56.5, 59.7] | 0.650 [0.619, 0.690] |
| W4 | 1132 | 1.71 (2.39) | 45.3 | 0.672 [0.655, 0.691] | 42.8 [40.5, 45.0] | 66.9 [65.0, 68.9] | 0.865 [0.815, 0.922] |
| W5 | 1038 | 2.04 (2.57) | 41.5 | 0.639 [0.608, 0.670] | 38.6 [36.4, 41.7] | 62.9 [59.9, 66.6] | 0.770 [0.696, 0.858] |

\* 95% confidence intervals in brackets; based on nonparametric cluster bootstrap with 1,000 replications, resampling villages with replacement to account for clustering of participants within villages (68)

The Theil decomposition was done for each survey wave separately, but there were minimal differences across waves (**Appendix Table S2**). Therefore, we focused on averages across the entire study period. Across nine stratifying variables, we found that between-group differences explained a remarkably small fraction of the observed concentration of stigma (**Figure 2**). For public stigma, between-group variation ranged from 0.12% (sex) to 2.6% (HIV), indicating that nearly all of the inequality in stigmatizing attitudes occurred within population subgroups rather than between them. The decomposition analysis for perceived stigma yielded similar findings.

**Figure 2.**
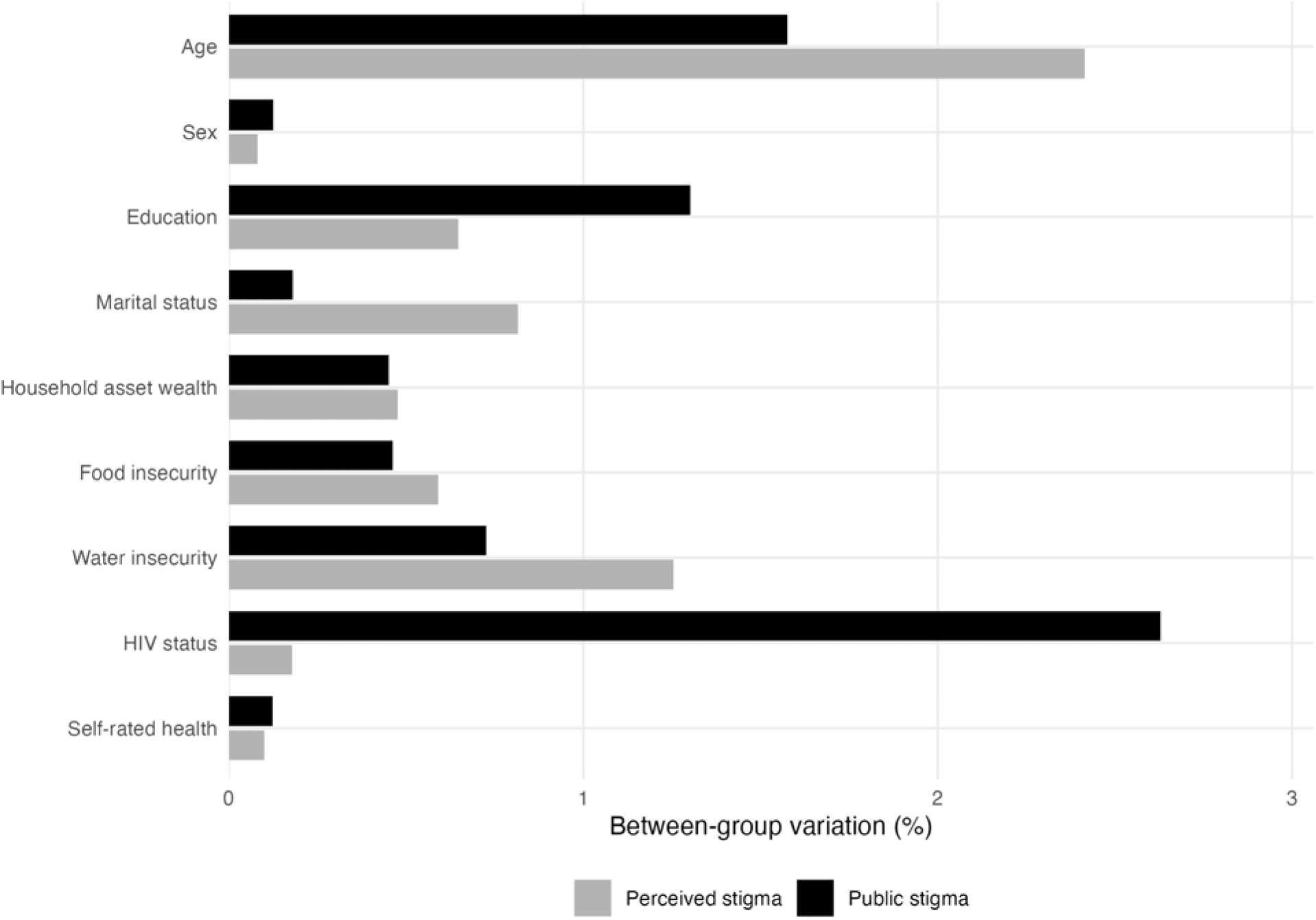
Theil decomposition, averaged across all 5 survey waves.

### Longitudinal analyses

The cohort design revealed fundamentally different dynamics underlying the declines in public vs. perceived stigma. In analyses restricted to the balanced panel of individuals participating in all five survey waves (N=869), public stigma showed meaningful between-person variance, with an ICC of 0.27 (95% CI, 0.23-0.30) and a detrended ICC of 0.35 (95% CI, 0.31-0.38) (**Appendix Table S3**). In contrast, perceived stigma showed essentially no between-person variance, with an ICC indistinguishable from zero.

Consistent with the ICC findings, public stigma showed moderate rank-order stability across survey waves (**Figure 3**). Across the study period, the mean correlation between waves was r=0.38; the correlation between baseline and endline was r=0.41 (95% CI, 0.34-0.47), indicating that study participants who had high levels of public stigma at baseline tended to have high levels of public stigma a decade later. Perceived stigma, in contrast, showed no rank-order stability, with an adjacent-wave mean correlation of r=0.08.

**Figure 3.**
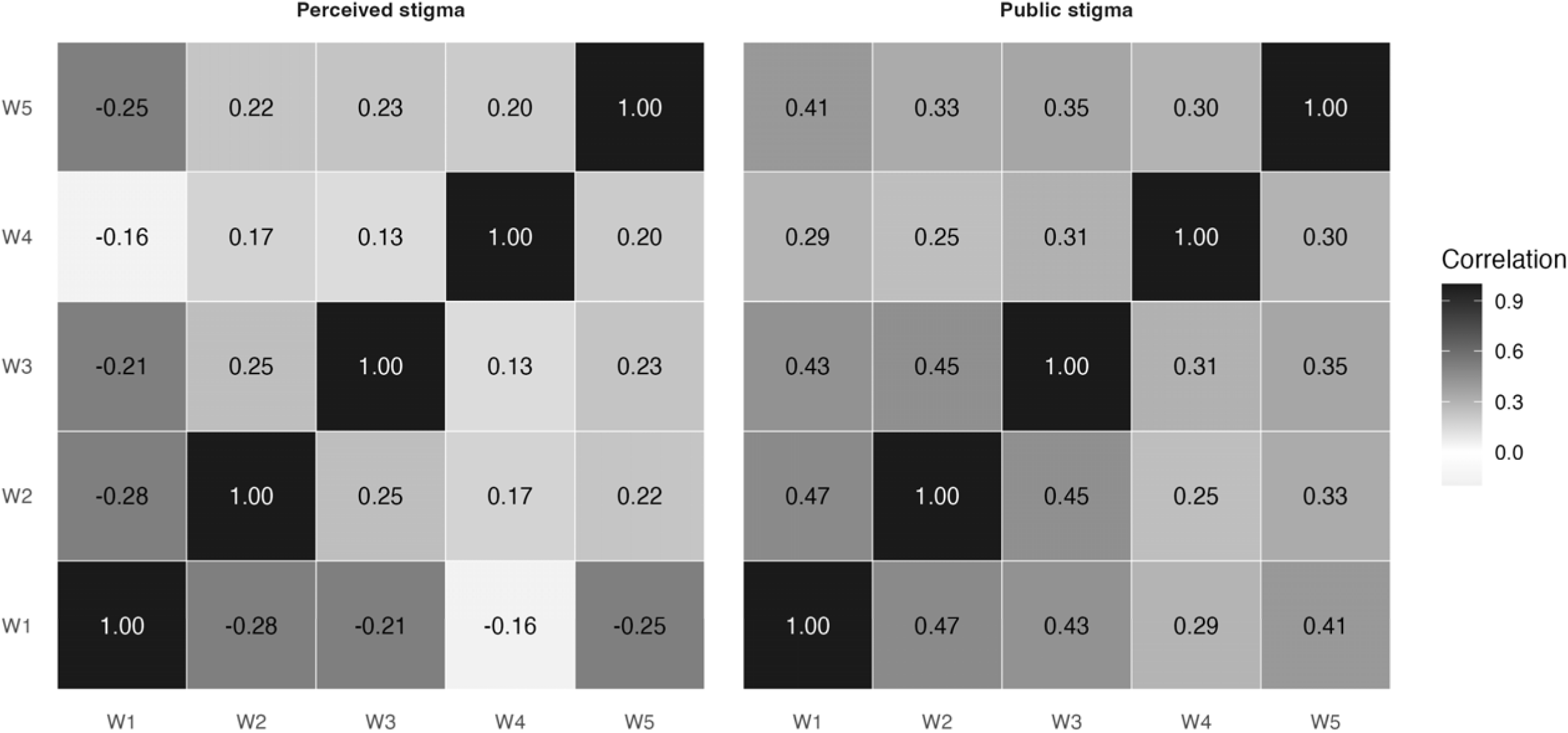
Rank-order stability across survey waves, 2014-24.

### Sensitivity analyses

For public stigma, the e-value analysis indicated that an omitted variable (e.g., social desirability bias) would need to have a strong association with both time and stigma (exceeding 3 on the relative risk ratio scale) to fully explain the declining trend in stigma. For perceived stigma, the e-value was even higher (8.3).

We compared the characteristics of the 869 retained study participants against the 907 study participants who were not retained (**Appendix Table S4**). Those who remained in the cohort at endline tended to be older; were more likely to be HIV positive and water insecure; and had fewer relationship disruptions, less formal education, and less public stigma (but more perceived stigma).

When we restricted estimation of the inequality measures to the balanced panel of 869 individuals who participated in all five survey waves, the balanced panel exhibited nearly identical trends in declining average stigma and increasing concentration of stigma over time (**Appendix Figure S3** and **Appendix Table S5**). The consistency of these findings suggests that the observed increasing concentration in stigma reflects true changes in stigma rather than compositional shifts.

Finally, to address the possibility of selection bias driven by differential attrition, we conducted IPC-weighted analyses, incorporating an additional 10 baseline health, psychosocial, and economic variables to predict retention through endline (**Appendix Table S6**). We re-estimated ICCs and rank-order stability correlations after applying these IPC weights. The IPC-weighted ICC and detrended ICC increased for both public stigma and perceived stigma but qualitatively the patterns remained unchanged (**Appendix Table S7**). The pattern of cross-wave correlations was also largely similar (**Appendix Figure S4**).

## DISCUSSION

In this population-based cohort study spanning 2014-2024 in rural Uganda, we documented substantial declines in both public HIV stigma (negative attitudes toward people with HIV) and perceived HIV stigma (perceptions about negative attitudes held by others). However, these aggregate trends masked an important distributional shift: stigma became markedly concentrated in a smaller subgroup of the population, with all measures of inequality rising over the study period. Decomposition of the Theil index specifically failed to identify any stratifying variables that explained an appreciable amount of this variation: nearly all the inequality in HIV stigma occurred within population subgroups rather than between them. In longitudinal analyses, public stigma demonstrated a higher degree of persistence than perceived stigma, indicating that personal stigmatizing attitudes remained relatively stable while perceptions of community norms shifted uniformly toward the floor.

The substantial decline in mean stigma observed in this study is consistent with broader trends documented in studies conducted in other African settings (9, 11, 29, 30, 32). The decline in perceived stigma was both steeper and more uniform than the decline in public stigma. At baseline, the mean perceived HIV stigma score of 8.8 indicates that study participants perceived that stigmatizing beliefs were the normative majority position across most domains. The endline mean of 2.4 indicated that the population went from believing “most people in my village hold most of these stigmatizing beliefs” to believing “most people in my village hold only a few of these stigmatizing beliefs.” This apparent resolution of pluralistic ignorance may itself facilitate further stigma reduction, as individuals who come to accurately recognize that most community members do not hold stigmatizing views may feel more comfortable expressing non-stigmatizing attitudes themselves (71).

Several mechanisms likely contributed to these trends. Individually targeted strategies such as information provision, contact interventions, and counseling have had mixed success in reducing stigma in the short term (3, 21–23). The widespread availability of effective treatment (9, 24–26) and the transformation of HIV from a fatal disease to a manageable chronic condition (72, 73) likely played a role. It is also possible that increasing education and anti-stigma campaigns may have played a role, but the population-level evidence supporting their efficacy is mixed (74, 75).

What our findings newly add to this body of literature is to suggest that, while these innovations may have been broadly effective in reducing HIV stigma broadly in the population, a minority subgroup did not respond. Borrowing inequality metrics widely used in the economics literature, we found that the “burden” of stigma became more unequally distributed in the population over time. The top 10% held approximately one-fourth of the total community burden of public stigma at baseline, and this had increased to more than one-third at endline; the top decile share of perceived stigma more than doubled during the study period. For both public stigma and perceived stigma, the Gini increased more slowly than the Theil, because stigma was becoming more concentrated in the upper tail of the distribution. These changes were also more apparent for perceived stigma (compared with public stigma), suggesting that perceptions of community norms shifted more rapidly than individuals’ negative attitudes.

A second key contribution of our study lies in the longitudinal analyses uniquely enabled by the cohort design (and not permitted by the serial cross-sectional designs that predominate in this literature). These analyses revealed fundamentally different dynamics underlying the declines in public vs. perceived stigma. Namely, public stigma demonstrated moderate rank-order stability over the study period, with an ICC that suggested trait-like properties. Perceived stigma, on the other hand, exhibited substantially different dynamics, with an ICC near zero (indicating essentially no stable individual differences) and universal convergence to low levels by the end of the study period. When the normative climate shifted (i.e., perhaps in response to scale-up of effective HIV treatment), most community members updated their perceptions collectively.

The persistence of a minority subgroup of individuals who retain stigmatizing beliefs, despite dramatic reductions in perceived stigma, has practical implications for programmatic interventions. These individuals apparently maintained their stigmatizing attitudes even as they increasingly recognized that such attitudes were no longer normative in their villages. This disconnect between personal attitudes and perceived norms suggests that different intervention approaches may be needed for these individuals. Further, decomposition of the Theil index showed that less than 3% of the inequality in HIV stigma was explained by demographic characteristics or variables related to economic status, psychosocial wellbeing, or health—suggesting that targeting of interventions will need to be based on other considerations.

### Limitations

Interpretation of our findings is subject to several important limitations. First, the study was conducted in a single rural area of Uganda. While Nyakabare Parish is typical of rural settings in Uganda, our findings may not generalize to urban settings in Uganda, or to either rural or urban settings outside of east Africa.

Second, our findings could have been affected by social desirability bias (7). Uganda has been the focus of numerous HIV prevention campaigns (76, 77). These could have prompted participants to focus on knowing what they *should* believe about people with HIV (and reporting accordingly) rather than changing what they *actually* believe, e.g., a form of “stereotype suppression” (78). This bias likely would have caused us to estimate a steeper decline in public stigma than had actually occurred. However, the e-values ranged from 4-7 indicating that there would need to be a high degree of confounding by social desirability bias. In previously published studies conducted in myriad settings, social desirability scores have demonstrated correlations with HIV stigma scales far below this threshold (79–81). Moreover, social desirability would have exerted less influence on perceived stigma—which is the opposite of what we found (i.e., if social desirability bias had been an important factor, the decline in public stigma should have been steeper than the decline in perceived stigma).

Related to the above, our measure of perceived HIV stigma could have been measured with error. Collective norms exist at multiple social levels (82, 83). It is therefore possible that specifying “other people in my village” as the reference group (rather than, for example, “other women my age”) may have resulted in our measuring the construct of perceived stigma with error.

Fourth, while the population sample size was relatively uniform from year to year, approximately half of baseline participants were lost to follow-up by endline, raising concerns about selective attrition. However, the consistency of results between the full sample and the balanced panel suggests that attrition did not substantially bias our findings. IPC weighted analyses did not substantively change our conclusions. The baseline survey had a more than 90% participation rate, and the IPC model included a rich set of demographic, psychosocial, and economic variables.

### Conclusion

In this decade-long population cohort study from rural Uganda, we observed substantial reductions in HIV stigma overall, including both public stigma (negative attitudes toward people living with HIV) and perceived stigma (beliefs about others’ negative attitudes). These average declines, which have been noted in previously published serial cross-sectional studies, obscured the fact that stigma became increasingly concentrated within a smaller segment of the population. When we decomposed one measure of inequality, we found that this persistent minority could not be identified by any measured stratifying characteristic; nearly all stigma-related inequality was attributable to differences within subgroups rather than differences between them. Longitudinally, public stigma showed greater persistence than perceived stigma, suggesting that individuals’ stigmatizing beliefs were comparatively stable, while perceived community norms declined more uniformly toward very low levels. Both sets of findings uniquely demonstrate the value of cohort designs for understanding stigma dynamics while also highlighting that intervention targeting will need to rely on methods other than sociodemographic profiling.

## Data Availability

Data are not available to be shared.

## REFERENCES

1. Nyblade L, Mingkwan P, Stockton MA. Stigma reduction: an essential ingredient to ending AIDS by 2030. Lancet HIV 2021;8(2):e106–e113.

2. Vella S, Wilson D. From Durban to Durban: end of AIDS further than hoped. Lancet HIV 2016;3(9):e403–e405.

3. Stangl AL, Lloyd JK, Brady LM, Holland CE, Baral S. A systematic review of interventions to reduce HIV-related stigma and discrimination from 2002 to 2013: how far have we come? J Int AIDS Soc 2013;16(3 Suppl 2):18734.

4. Kelly JA, St Lawrence JS, Hood HV, Smith S, Jr., Cook DJ. Nurses’ attitudes towards AIDS. J Contin Educ Nurs 1988;19(2):78–83.

5. Herek GM, Capitanio JP. Public reactions to AIDS in the United States: a second decade of stigma. Am J Public Health 1993;83(4):574–577.

6. Kalichman SC, Simbayi LC. HIV testing attitudes, AIDS stigma, and voluntary HIV counselling and testing in a black township in Cape Town, South Africa. Sex Transm Infect 2003;79(6):442–447.

7. Nyblade L, MacQuarrie K, Phillip F, Kwesigabo G, Mbwambo J, Ndega J, et al. *Measuring HIV stigma: results of a field test in Tanzania*. Washington, D.C.: U.S. Agency for International Development; 2005.

8. Wolfe WR, Weiser SD, Leiter K, Steward WT, Percy-de Korte F, Phaladze N, et al. The impact of universal access to antiretroviral therapy on HIV stigma in Botswana. Am J Public Health 2008;98(10):1865–1871.

9. Chan BT, Tsai AC. HIV stigma trends in the general population during antiretroviral treatment expansion: analysis of 31 countries in sub-Saharan Africa, 2003-2013. J Acquir Immune Defic Syndr 2016;72(5):558–564.

10. Hargreaves JR, Krishnaratne S, Mathema H, Lilleston PS, Sievwright K, Mandla N, et al. Individual and community-level risk factors for HIV stigma in 21 Zambian and South African communities: analysis of data from the HPTN071 (PopART) study. AIDS 2018;32(6):783–793.

11. Doyle CM, Kuchukhidze S, Stannah J, Flores Anato JL, Xia Y, Logie CH, et al. The impact of HIV stigma and discrimination on HIV testing, antiretroviral treatment, and viral suppression in Africa: a pooled analysis of population-based surveys. Lancet HIV 2026;13(4):e235–e246.

12. Kalichman SC, Shkembi B, Wanyenze RK, Naigino R, Bateganya MH, Menzies NA, et al. Perceived HIV stigma and HIV testing among men and women in rural Uganda: a population-based study. Lancet HIV 2020;7(12):e817–e824.

13. Jones HS, Floyd S, Stangl A, Bond V, Hoddinott G, Pliakas T, et al. Association between HIV stigma and antiretroviral therapy adherence among adults living with HIV: baseline findings from the HPTN 071 (PopART) trial in Zambia and South Africa. Trop Med Int Health 2020;25(10):1246–1260.

14. Hargreaves JR, Pliakas T, Hoddinott G, Mainga T, Mubekapi-Musadaidzwa C, Donnell D, et al. HIV stigma and viral suppression among people living with HIV in the context of universal test and treat: analysis of data from the HPTN 071 (PopART) trial in Zambia and South Africa. J Acquir Immune Defic Syndr 2020;85(5):561–570.

15. Katz IT, Ryu AE, Onuegbu AG, Psaros C, Weiser SD, Bangsberg DR, et al. Impact of HIV-related stigma on treatment adherence: systematic review and meta-synthesis. J Int AIDS Soc 2013;16(Suppl 2):18640.

16. Takada S, Nyakato V, Nishi A, O’Malley AJ, Kakuhikire B, Perkins JM, et al. The social network context of HIV stigma: Population-based, sociocentric network study in rural Uganda. Soc Sci Med 2019;233:229–236.

17. Takada S, Weiser SD, Kumbakumba E, Muzoora C, Martin JN, Hunt PW, et al. The dynamic relationship between social support and HIV stigma in rural Uganda. Ann Behav Med 2014;48(1):26–37.

18. Ware NC, Idoko J, Kaaya S, Biraro IA, Wyatt MA, Agbaji O, et al. Explaining adherence success in sub-Saharan Africa: an ethnographic study. PLoS Med 2009;6(1):e11.

19. Tsai AC, Bangsberg DR, Frongillo EA, Hunt PW, Muzoora C, Martin JN, et al. Food insecurity, depression and the modifying role of social support among people living with HIV/AIDS in rural Uganda. Soc Sci Med 2012;74(12):2012–2019.

20. Yu H. Social stigma as a barrier to HIV testing: Evidence from a randomized experiment in Mozambique. J Dev Econ 2023;161(Mar):103035.

21. Sengupta S, Banks B, Jonas D, Miles MS, Smith GC. HIV interventions to reduce HIV/AIDS stigma: a systematic review. AIDS Behav 2011;15(6):1075–1087.

22. Mak WWS, Mo PKH, Ma GYK, Lam MYY. Meta-analysis and systematic review of studies on the effectiveness of HIV stigma reduction programs. Soc Sci Med 2017;188:30–40.

23. Ferguson L, Gruskin S, Bolshakova M, Rozelle M, Yagyu S, Kasoka K, et al. Systematic review and quantitative and qualitative comparative analysis of interventions to address HIV-related stigma and discrimination. AIDS 2023;37(13):1919–1939.

24. Farmer P, Leandre F, Mukherjee JS, Claude M, Nevil P, Smith-Fawzi MC, et al. Community-based approaches to HIV treatment in resource-poor settings. Lancet 2001;358(9279):404–409.

25. Castro A, Farmer P. Understanding and addressing AIDS-related stigma: from anthropological theory to clinical practice in Haiti. Am J Public Health 2005;95(1):53–59.

26. Chan BT, Tsai AC, Siedner MJ. HIV treatment scale-up and HIV-related stigma in sub-Saharan Africa: a longitudinal cross-country analysis. Am J Pub Health 2015;105(8):1581–1587.

27. Stangl AL, Pliakas T, Mainga T, Steinhaus M, Mubekapi-Musadaidzwa C, Viljoen L, et al. The effect of universal testing and treatment on HIV stigma in 21 communities in Zambia and South Africa. AIDS 2020;34(14):2125–2135.

28. Viljoen L, Bond VA, Reynolds LJ, Mubekapi-Musadaidzwa C, Baloyi D, Ndubani R, et al. Universal HIV testing and treatment and HIV stigma reduction: a comparative thematic analysis of qualitative data from the HPTN 071 (PopART) trial in South Africa and Zambia. Sociol Health Illn 2021;43(1):167–185.

29. Mall S, Middelkoop K, Mark D, Wood R, Bekker LG. Changing patterns in HIV/AIDS stigma and uptake of voluntary counselling and testing services: the results of two consecutive community surveys conducted in the Western Cape, South Africa. AIDS Care 2013;25(2):194–201.

30. Nuwaha F, Kasasa S, Wana G, Muganzi E, Tumwesigye E. Effect of home-based HIV counselling and testing on stigma and risky sexual behaviours: serial cross-sectional studies in Uganda. J Int AIDS Soc 2012;15(2):17423.

31. Tam G, Wong NS, Lee SS. Serial surveys of Hong Kong medical students regarding attitudes towards HIV/AIDS from 2007 to 2017. Hong Kong Med J 2022;28(3):223–229.

32. Visser MJ. Change in HIV-related stigma in South Africa between 2004 and 2016: a cross-sectional community study. AIDS Care 2018;30(6):734–738.

33. Maughan-Brown B. Stigma rises despite antiretroviral roll-out: a longitudinal analysis in South Africa. Soc Sci Med 2010;70(3):368–374.

34. Bebell LM, Kembabazi A, Musinguzi N, Martin JN, Hunt PW, Boum Y, 2nd, et al. Internalized stigma, depressive symptoms, and the modifying role of antiretroviral therapy: A cohort study in rural Uganda. SSM Ment Health 2021;1:100034.

35. Tsai AC, Bangsberg DR, Kegeles SM, Katz IT, Haberer JE, Muzoora C, et al. Internalized stigma, social distance, and disclosure of HIV seropositivity in rural Uganda. Ann Behav Med 2013;46(3):285–294.

36. Tsai AC, Bangsberg DR, Bwana M, Haberer JE, Frongillo EA, Muzoora C, et al. How does antiretroviral treatment attenuate the stigma of HIV? Evidence from a cohort study in rural Uganda. AIDS Behav 2013;17(8):2725–2731.

37. Logie CH, Marcus N, Wang Y, Kaida A, O’Campo P, Ahmed U, et al. A longitudinal study of associations between HIV-related stigma, recent violence and depression among women living with HIV in a Canadian cohort study. J Int AIDS Soc 2019;22(7):e25341.

38. Norcini-Pala A, Stringer KL, Kempf MC, Konkle-Parker D, Wilson TE, Tien PC, et al. Longitudinal associations between intersectional stigmas, antiretroviral therapy adherence, and viral load among women living with HIV using multidimensional latent transition item response analysis. Soc Sci Med 2025;366:117643.

39. Uganda Bureau of Statistics. National population and housing census 2024: final report, vol. 1. Kampala: Uganda Bureau of Statistics; 2024.

40. Perkins JM, Nyakato VN, Kakuhikire B, Tsai AC, Subramanian SV, Bangsberg DR, et al. Food insecurity, social networks and symptoms of depression among men and women in rural Uganda: a cross-sectional, population-based study. Public Health Nutr 2018;21(5):838–848.

41. Mushavi RC, Burns BFO, Kakuhikire B, Owembabazi M, Vořechovská D, McDonough AQ, et al. “When you have no water, it means you have no peace”: A mixed-methods, whole-population study of water insecurity and depression in rural Uganda. Soc Sci Med 2020;245:112561.

42. Uganda Ministry of Health. Uganda Population-Based HIV Impact Assessment (UPHIA) 2016-17. Kampala: Uganda Ministry of Health; 2019.

43. Uganda Ministry of Health. Uganda clinical guidelines 2016: national guidelines for management of common conditions. Kampala: Ministry of Health Uganda; 2016.

44. Dirlikov E, Kamoga J, Talisuna SA, Namusobya J, Kasozi DE, Akao J, et al. Scale-up of HIV antiretroviral therapy and estimation of averted infections and HIV-related deaths – Uganda, 2004-2022. MMWR Morb Mortal Wkly Rep 2023;72(4):90–94.

45. Tsai AC, Kakuhikire B, Perkins JM, Downey JM, Baguma C, Satinsky EN, et al. Normative vs personal attitudes toward persons with HIV, and the mediating role of perceived HIV stigma in rural Uganda. J Glob Health 2021;11:04956.

46. Kakuhikire B, Satinsky EN, Baguma C, Rasmussen JD, Perkins JM, Gumisiriza P, et al. Correlates of attendance at community engagement meetings held in advance of bio-behavioral research studies: A longitudinal, sociocentric social network study in rural Uganda. PLoS Med 2021;18(7):e1003705.

47. Corrigan PW, Watson AC. The paradox of self-stigma and mental illness. Clin Psychol Sci Pract 2002;9(1):35–53.

48. Pryor JB, Reeder GD, Vinacco R, Kott TL. The instrumental and symbolic functions of attitudes towards persons with AIDS. J Appl Soc Psychol 1989;19(5):377–404.

49. Tsai AC, Bangsberg DR, Weiser SD. Harnessing poverty alleviation to reduce the stigma of HIV in sub-Saharan Africa. PLoS Med 2013;10(11):e1001557.

50. Tsai AC, Hatcher AM, Bukusi EA, Weke E, Lemus Hufstedler L, Dworkin SL, et al. A livelihood intervention to reduce the stigma of HIV in rural Kenya: longitudinal qualitative study. AIDS Behav 2017;21(1):248–260.

51. Filmer D, Pritchett LH. Estimating wealth effects without expenditure data –– or tears: an application to educational enrollments in states of India. Demography 2001;38(1):115–132.

52. Smith ML, Kakuhikire B, Baguma C, Rasmussen JD, Bangsberg DR, Tsai AC. Do household asset wealth measurements depend on who is surveyed? Asset reporting concordance within multi-adult households in rural Uganda. J Glob Health 2020;10(1):010412.

53. Ballard T, Coates J, Swindale A, Deitchler M. *Household Hunger Scale: indicator definition and measurement guide*. Washington, D.C.: Food and Nutrition Technical Assistance III Project; 2011.

54. Tsai AC, Bangsberg DR, Emenyonu N, Senkungu JK, Martin JN, Weiser SD. The social context of food insecurity among persons living with HIV/AIDS in rural Uganda. Soc Sci Med 2011;73(12):1717–1724.

55. Swindale A, Bilinsky P. Development of a universally applicable household food insecurity measurement tool: process, current status, and outstanding issues. J Nutr 2006;136(5):1449S–1452S.

56. Tsai AC, Kakuhikire B, Mushavi R, Vořechovská D, Perkins JM, McDonough AQ, et al. Population-based study of intra-household gender differences in water insecurity: reliability and validity of a survey instrument for use in rural Uganda. J Water Health 2016;14(2):280–292.

57. Idler EL, Benyamini Y. Self-rated health and mortality: a review of twenty-seven community studies. J Health Soc Behav 1997;38(1):21–37.

58. Wagstaff A, Paci P, van Doorslaer E. On the measurement of inequalities in health. Soc Sci Med 1991;33(5):545–557.

59. Theil H. Economics and information theory. Studies in Mathematical and Managerial Economics, Vol. 7. Amsterdam: North-Holland; 1967.

60. Cowell FA, Flachaire E. Income distribution and inequality measurement: The problem of extreme values. J Econometrics 2007;141(2):1044–1072.

61. Atkinson AB, Piketty T, Saez E. Top incomes in the long run of history. J Econ Lit 2011;49(1):3–71.

62. Piketty T, Saez E. Income inequality in the United States, 1913-1998. Q J Econ 2003;118(1):1–41.

63. Pradhan M, Sahn DE, Younger SD. Decomposing world health inequality. J Health Econ 2003;22(2):271–293.

64. Gakidou EE, Murray CJ, Frenk J. Defining and measuring health inequality: an approach based on the distribution of health expectancy. Bull World Health Organ 2000;78(1):42–54.

65. Shorrocks AF. The class of additively decomposable inequality measures. Econometrica 1980;48(3):613–625.

66. Shorrocks AF. Inequality decomposition by population subgroups. Econometrica 1984;52(6):1369–1385.

67. VanderWeele TJ, Ding P. Sensitivity analysis in observational research: introducing the E-value. Ann Intern Med 2017;167(4):268–274.

68. Cameron AC, Gelbach JB, Miller DL. Bootstrap-based improvements for inference with clustered errors. Rev Econ Stat 2008;90(3):414–427.

69. Bell RM, McCaffrey DF. Bias reduction in standard errors for linear regression with multi-stage samples. Surv Methodol 2002;28(2):169–181.

70. Pustejovsky JE, Tipton P. Small-sample methods for clusterrobust variance estimation and hypothesis testing in fixed effects models. J Bus Econ Stat 2018;36(4):672–683.

71. Prentice DA, Miller DT. Pluralistic ignorance and alcohol use on campus: some consequences of misperceiving the social norm. J Pers Soc Psychol 1993;64(2):243–256.

72. Mills EJ, Bakanda C, Birungi J, Chan K, Ford N, Cooper CL, et al. Life expectancy of persons receiving combination antiretroviral therapy in low-income countries: a cohort analysis from Uganda. Ann Intern Med 2011;155(4):209–216.

73. Bor J, Herbst AJ, Newell ML, Barnighausen T. Increases in adult life expectancy in rural South Africa: valuing the scale-up of HIV treatment. Science 2013;339(6122):961–965.

74. Tsai AC, Venkataramani AS. The causal effect of education on HIV stigma in Uganda: evidence from a natural experiment. Soc Sci Med 2015;142(1):37–46.

75. Yang D, Allen Jt, Mahumane A, Riddell Jt, Yu H. Knowledge, stigma, and HIV testing: an analysis of a widespread HIV/AIDS program. J Dev Econ 2023;160:102958.

76. Murphy EM, Greene ME, Mihailovic A, Olupot-Olupot P. Was the “ABC” approach (abstinence, being faithful, using condoms) responsible for Uganda’s decline in HIV? PLoS Med 2006;3(9):e379.

77. de Walque D. How does the impact of an HIV/AIDS information campaign vary with educational attainment? Evidence from rural Uganda. J Dev Econ 2007;84(2):686–714.

78. Corrigan PW, Penn DL. Lessons from social psychology on discrediting psychiatric stigma. Am Psychol 1999;54(9):765–776.

79. Wagner AC, Hart TA, McShane KE, Margolese S, Girard TA. Health care provider attitudes and beliefs about people living with HIV: Initial validation of the Health Care Provider HIV/AIDS Stigma Scale (HPASS). AIDS Behav 2014;18(12):2397–2408.

80. Kipp AM, Audet CM, Earnshaw VA, Owens J, McGowan CC, Wallston KA. Re-validation of the Van Rie HIV/AIDS-related stigma scale for use with people living with HIV in the United States. PLoS One 2015;10(3):e0118836.

81. Cahill L, Gifford AJ, Jones BA, McDermott DT. The HIV Anxiety Scale (HAS): developing and validating a measure of Human Immunodeficiency Virus (HIV) anxiety. AIDS Behav 2025;29(7):2258–2271.

82. Borsari B, Carey KB. Descriptive and injunctive norms in college drinking: a meta-analytic integration. J Stud Alcohol 2003;64(3):331–341.

83. Lapinski MK, Rimal RN. An explication of social norms. Comm Theory 2005;15(2):127–147.

