## Appendix for "Declining but increasingly concentrated HIV stigma in rural Uganda: population-based cohort study, 2014-2024"

*Contents*

**Text S1.** Methods

**Table S1.** Characteristics of the study population, by survey wave

**Table S2.** Theil decomposition, by survey wave

**Table S3.** Unconditional and detrended intraclass correlation coefficients for public stigma and perceived stigma

**Table S4.** Characteristics of the study population, comparing those who were retained in all five survey waves (N=869) vs. those who were not (N=907)

**Table S5.** Trends in mean stigma and concentration, comparing estimates from the full serial cross sections vs. balanced panel only

**Table S6.** Correlates of retention in all five survey waves

**Table S7.** Unconditional and detrended intraclass correlation coefficients for public stigma and perceived stigma, under progressive trimming of inverse probability of censoring weights

**Figure S1.** Flow diagram depicting the open cohort design

**Figure S2.** Distribution of HIV stigma scores, by survey wave

**Figure S3.** Trends in mean stigma and concentration, comparing estimates from the full serial cross sections vs. balanced panel only

**Figure S4.** Rank-order stability across survey waves, after application of inverse probability of censoring weights

**Text S1. Methods**

*Ethics approval.* All research assistants underwent intensive training in fieldwork procedures and survey administration for collecting sensitive information, including guidance on how to pause the interview if another person came within earshot of the interview. They also received two supplementary trainings—one from The AIDS Support Organization and one from a Ugandan counseling psychologist—focused on responding to sensitive disclosures from study participants. Each eligible individual was approached in the field, usually at their home or workplace, by a research assistant fluent in the local language (Runyankore). The research assistant briefly presented the study as a community survey about the social lives and health of residents of Nyakabare Parish (i.e., not as a study specifically about attitudes toward people with HIV) and invited them to take part. For individuals who indicated possible interest, the study was then explained in full, and written informed consent was obtained. Participants who were unable to write were allowed to provide consent via thumbprint.

We sought input on the study design from a community advisory board consisting of eight community leaders (four women and four men), including the district community development officer and an HIV-positive volunteer counselor from the Mbarara Regional Referral Hospital Immune Suppression Syndrome Clinic. In addition, we held a series of community sensitization meetings in each study village, during which we described the study design in detail, responded to questions, and invited feedback (Kakuhikire et al., 2021).

*Data collection during COVID-19.* Data collection was temporarily halted on March 20, 2020, during the third survey wave, due to country-wide restrictions on internal movement implemented to suppress the spread of COVID-19. Data collection resumed April 21; the Mbarara University of Science and Technology Research Ethics Committee and the Partners Human Research Committee, consistent with guidance from the Uganda National Council of Science and Technology, authorized a modification from written informed consent and in-person interview procedures to verbal informed consent and mobile phone interviews. In-person interviews resumed on July 26, 2022, near the end of the fourth survey wave.

The censoring model included the 11 variables from Table 1 plus 10 additional covariates. We asked participants to report the number of days of work they missed due to illness or injury in the past month, the number of community groups to which they belonged (Kakuhikire et al., 2021), and the size of their emotional support network (Burt, 1984). Waist circumference measurements were taken at the level of the iliac crest (U.S. National Center for Health Statistics, 1996), with obesity defined as a waist circumference measure >102 cm among men and >88 cm among women (NHLBI Obesity Education Initiative Expert Panel on the Identification, 1998). Heavy alcohol use was measured using the 3-item scale developed in Mbarara by Fatch et al. (2013). We measured cognitive social capital and perceived trust using 2-item scales from Pronyk et al. (2006). To elicit study participants’ beliefs in the justifiability of intimate partner violence, we administered five questions modeled after those included in the Demographic and Health Surveys (Kishor & Johnson, 2004; Tsai et al., 2017). Depressive symptom severity was measured using a modified version of the Hopkins Symptom Checklist for Depression (Bolton & Ndogoni, 2001; Mushavi et al., 2020). We measured happiness with a single-item scale modeled after the single-item measure included in the U.S. General Social Survey (Andrews & Withey, 1976; Gurin, Veroff, & Feld, 1960).

We fitted a logistic regression model specifying retention in all survey five waves as the dependent variable and the 21 covariates listed above (**Appendix Table S6**). We calculated stabilized IPC weights (Cole & Hernán, 2008; Hernán & Robins, 2006) as the marginal retention probability divided by each individual’s predicted probability of retention, then rescaled to a mean of 1. The IPC weights upweight observations from participants with characteristics similar to those lost to follow-up, effectively creating a pseudo-population representative of the original baseline sample (Robins, Hernán, & Brumback, 2000). We applied the IPC weights to re-estimate ICCs and rank-order stability correlations. To assess the influence of extreme weights, we repeated the IPC-weighted analyses after trimming weights at the 1st/99th, 5th/95th, and 10th/90th percentiles. The IPC-weighted ICC and detrended ICC increased for both public stigma and perceived stigma but qualitatively the patterns remained unchanged (**Appendix Table S7**). The pattern of cross-wave correlations was also largely similar (**Appendix Figure S4**).

*Limitations.* Interpretation of our findings is subject to several important limitations. First, the study was conducted in a single rural area of Uganda. While Nyakabare Parish is typical of rural settings in Uganda, our findings may not generalize to urban settings in Uganda, or to either rural or urban settings outside of east Africa.

Second, our findings could have been affected by social desirability bias (Nyblade et al., 2005). Uganda has been the focus of numerous HIV prevention campaigns (de Walque, 2007; Murphy et al., 2006). These could have prompted participants to focus on knowing what they *should* believe about people with HIV (and reporting accordingly) rather than changing what they *actually* believe, e.g., a form of “stereotype suppression” (Corrigan & Penn, 1999). This bias likely would have caused us to estimate a steeper decline in public stigma than had actually occurred. However, the e-values ranged from 4-7 indicating that there would need to be a high degree of confounding by social desirability bias. In previously published studies conducted in myriad settings, social desirability scores have demonstrated correlations with HIV stigma scales far below this threshold (Cahill et al., 2025; Kipp et al., 2015; Wagner et al., 2014). Moreover, social desirability would have exerted less influence on perceived stigma—which is the opposite of what we found (i.e., if social desirability bias had been an important factor, the decline in public stigma should have been steeper than the decline in perceived stigma).

Related to the above, our measure of perceived HIV stigma could have been measured with error. Collective norms exist at multiple social levels (Borsari & Carey, 2003; Lapinski & Rimal, 2005). It is therefore possible that specifying “other people in my village” as the reference group (rather than, for example, “other women my age”) may have resulted in our measuring the construct of perceived stigma with error.

Fourth, while the population sample size was relatively uniform from year to year, approximately half of baseline participants were lost to follow-up by endline, raising concerns about selective attrition. However, the consistency of results between the full sample and the balanced panel suggests that attrition did not substantially bias our findings. IPC weighted analyses did not substantively change our conclusions. The baseline survey had a more than 90% participation rate, and the IPC model included a rich set of demographic, psychosocial, and economic variables.

**Table S1.** Characteristics of the study population, by survey wave

| **Variable** | **W1** | **W2** | **W3** | **W4** | **W5** |
| --- | --- | --- | --- | --- | --- |
| N | 1776 | 1601 | 1603 | 1548 | 1687 |
| Age, mean (SD) | 38.1 (16.8) | 40.0 (16.7) | 41.9 (17.0) | 42.3 (16.0) | 41.9 (16.3) |
| Female, % | 54.6 | 55.4 | 54.8 | 55.1 | 53.2 |
| Education, % | |  |  |  |  |
| No formal education | 28.4 | 27.2 | 28.8 | 28 | 27.2 |
| Some primary | 22.4 | 24.6 | 23 | 21.2 | 22.6 |
| Completed primary | 35.3 | 36.2 | 35.9 | 39.1 | 40.5 |
| Secondary or more | 0 | 0 | 0 | 0 | 0 |
| Marital status, % | |  |  |  |  |
| Never married | 58.5 | 61.6 | 62.8 | 65.7 | 62.8 |
| Married/cohabitating | 17 | 17.1 | 17.6 | 17.5 | 18.6 |
| Separated/divorced/widowed | 24.5 | 21.4 | 19.7 | 16.8 | 18.6 |
| Household asset wealth, mean (SD) | 0.0 (2.1) | 0.0 (2.1) | -0.0 (2.1) | 0.0 (2.1) | 0.0 (2.0) |
| Household Hunger Scale, mean (SD) | 0.5 (1.0) | 0.5 (1.5) | 0.3 (1.1) | 0.2 (1.0) | 0.4 (1.3) |
| Household Water Insecurity Access Scale, mean (SD) | 7.7 (6.8) | 3.8 (5.3) | 2.4 (4.6) | 1.8 (4.0) | 2.3 (4.4) |
| Self-rated health, % | |  |  |  |  |
| Poor | 1.1 | 0.7 | 1 | 1.2 | 0.8 |
| Fair | 84.8 | 83.1 | 77.7 | 81.8 | 77.8 |
| Good | 14.2 | 16.2 | 21.3 | 16.9 | 21.4 |
| Excellent | 0 | 0 | 0 | 0 | 0 |
| HIV positive by self-report, % | 8.7 | 10.4 | 11.4 | 11.6 | 12.3 |
| Public stigma score, mean (SD) | 3.7 (3.2) | 2.2 (2.1) | 1.8 (2.0) | 1.2 (1.7) | 1.5 (1.8) |
| Perceived stigma score, mean (SD) | 8.8 (3.9) | 3.7 (3.3) | 2.8 (3.2) | 1.8 (2.5) | 2.4 (2.9) |

**Table S2.** Theil decomposition, by survey wave

*Panel A. Public stigma*

| **Stratifier** | **W1** | **W2** | **W3** | **W4** | **W5** | **Average** |
| --- | --- | --- | --- | --- | --- | --- |
| Age | 1.95 | 1.85 | 1.46 | 0.51 | 2.12 | 1.58 |
| Sex | 0.26 | 0.11 | 0.04 | 0.21 | 0.01 | 0.12 |
| Education | 3.58 | 0.56 | 0.82 | 1.24 | 0.3 | 1.3 |
| Marital status | 0.27 | 0.13 | 0.12 | 0.04 | 0.34 | 0.18 |
| Household asset wealth | 1.3 | 0.15 | 0.54 | 0.09 | 0.17 | 0.45 |
| Food insecurity | 1.36 | 0.12 | 0.13 | 0.4 | 0.3 | 0.46 |
| Water insecurity | 0.42 | 0.85 | 0.51 | 1.24 | 0.62 | 0.73 |
| HIV status | 2.99 | 3.33 | 1.53 | 2.01 | 3.29 | 2.63 |
| Self-rated health | 0.11 | 0.05 | 0 | 0.45 | 0 | 0.12 |

*Panel B. Perceived stigma*

| **Stratifier** | **W1** | **W2** | **W3** | **W4** | **W5** | **Average** |
| --- | --- | --- | --- | --- | --- | --- |
| Age | 2.77 | 3.7 | 1.3 | 1.15 | 3.16 | 2.42 |
| Sex | 0.17 | 0.14 | 0.03 | 0 | 0.06 | 0.08 |
| Education | 1.68 | 0.58 | 0.21 | 0.08 | 0.68 | 0.65 |
| Marital status | 0.57 | 1.77 | 0.14 | 0.28 | 1.32 | 0.82 |
| Household asset wealth | 1.14 | 0.07 | 0.35 | 0.49 | 0.33 | 0.48 |
| Food insecurity | 0.28 | 0.35 | 0.54 | 1.55 | 0.24 | 0.59 |
| Water insecurity | 0.15 | 1.1 | 1.09 | 2.36 | 1.58 | 1.25 |
| HIV status | 0.26 | 0.17 | 0.21 | 0.03 | 0.21 | 0.18 |
| Self-rated health | 0.13 | 0.15 | 0.08 | 0.08 | 0.06 | 0.1 |

**Table S3.** Unconditional and detrended intraclass correlation coefficients for public stigma and perceived stigma

| **Measure** | **Individuals** | **Observations** | **ICC*** | **ICC (Detrended)*** |
| --- | --- | --- | --- | --- |
| Public stigma | 869 | 4345 | 0.26 [0.24, 0.28] | 0.34 [0.31, 0.37] |
| Perceived stigma | 869 | 4345 | 0.00 [0.00, 0.00] | 0.00 [0.00, 0.03] |

** 95% confidence intervals in brackets; based on nonparametric cluster bootstrap with 1,000 replications, resampling villages with replacement to account for clustering of participants within villages (68)*

**Table S4.** Characteristics of the study population, comparing those who were retained in all five survey waves (N=869) vs. those who were not (N=907)

| **Variable** | **Retained** | **Lost to Follow-up** | **P-value** |
| --- | --- | --- | --- |
| Age, mean (SD) | 40.21 (13.92) | 36.08 (18.95) | 0.003 |
| Female, n (%) | 480 (55.2%) | 489 (53.9%) | 0.658 |
| Education, n (%) | |  | <0.001 |
| No formal education | 291 (38.5%) | 214 (27.7%) |  |
| Some primary | 224 (29.6%) | 173 (22.4%) |  |
| Completed primary | 241 (31.9%) | 386 (49.9%) |  |
| Marital status, n (%) | |  | <0.001 |
| Never married | 624 (71.9%) | 413 (45.6%) |  |
| Married/cohabiting | 149 (17.2%) | 153 (16.9%) |  |
| Sep/Div/Widowed | 95 (10.9%) | 340 (37.5%) |  |
| Household asset wealth, mean (SD) | -0.05 (1.94) | 0.09 (2.25) | 0.333 |
| Household Hunger Scale, mean (SD) | 0.57 (1.07) | 0.50 (1.03) | 0.426 |
| Household Water Insecurity, mean (SD) | 8.40 (6.86) | 7.02 (6.57) | 0.064 |
| HIV positive, n (%) | 110 (12.7%) | 44 (4.9%) | <0.001 |
| Good self-rated health, n (%) | 111 (12.8%) | 140 (15.5%) | 0.269 |
| Baseline public stigma, mean (SD) | 3.54 (2.92) | 3.88 (3.36) | 0.023 |
| Baseline perceived stigma, mean (SD) | 9.26 (3.58) | 8.31 (4.17) | 0.008 |

**Table S5.** Trends in mean stigma and concentration, comparing estimates from the full serial cross sections vs. balanced panel only

|  |  |  | **Mean** | | | **Gini** | | |
| --- | --- | --- | --- | --- | --- | --- | --- | --- |
| **Sample** | **N (Baseline)** | **N (Endline)** | **Baseline** | **Endline** | **Change** | **Baseline** | **Endline** | **Change** |
| *Panel A. Public stigma* | |  |  |  |  |  |  |  |
| Full Sample | 1776 | 1687 | 3.71 | 1.46 | -2.25 | 0.46 | 0.614 | 0.154 |
| Balanced Panel | 869 | 869 | 3.54 | 1.36 | -2.17 | 0.446 | 0.642 | 0.195 |
| *Panel B. Perceived stigma* | |  |  |  |  |  |  |  |
| Full Sample | 1776 | 1687 | 8.78 | 2.37 | -6.4 | 0.252 | 0.624 | 0.372 |
| Balanced Panel | 869 | 869 | 9.26 | 2.01 | -7.25 | 0.216 | 0.643 | 0.427 |

**Table S6.** Correlates of retention in all five survey waves

| **Variable** | **OR** | **95% Confidence Interval*** | **P-value** |
| --- | --- | --- | --- |
| (Intercept) | 0.14 | (0.06, 0.34) | 0.004 |
| Age | 1.03 | (1.02, 1.05) | 0.007 |
| Female | 1.02 | (0.84, 1.24) | 0.855 |
| Education |  |  |  |
| No formal education | Ref |  |  |
| Some primary | 1.05 | (0.75, 1.48) | 0.789 |
| Completed primary | 0.69 | (0.54, 0.86) | 0.017 |
| Married/cohabiting | 0.72 | (0.40, 1.29) | 0.307 |
| Household asset wealth | 0.97 | (0.93, 1.01) | 0.238 |
| Household Hunger Scale | 1.03 | (0.86, 1.24) | 0.744 |
| Household Water Insecurity | 1.03 | (1.00, 1.06) | 0.105 |
| HIV positive | 2.68 | (2.06, 3.48) | <0.001 |
| Good self-rated health | 0.94 | (0.69, 1.28) | 0.693 |
| Public stigma score | 1.03 | (0.99, 1.08) | 0.200 |
| Perceived stigma score | 1.06 | (1.01, 1.10) | 0.040 |
| Days work missed | 1.00 | (0.97, 1.03) | 0.934 |
| Obese | 1.15 | (0.96, 1.37) | 0.172 |
| Heavy alcohol use | 1.19 | (0.86, 1.63) | 0.336 |
| Community group memberships | 1.3 | (1.18, 1.43) | 0.001 |
| Cognitive social capital | 1.21 | (0.74, 1.97) | 0.486 |
| Perceived trust | 0.99 | (0.84, 1.16) | 0.883 |
| IPV belief score | 0.98 | (0.92, 1.05) | 0.552 |
| Depressive symptoms | 0.72 | (0.61, 0.85) | 0.006 |
| Happiness | 0.85 | (0.66, 1.09) | 0.243 |
| Emotional support network size | 1.13 | (1.02, 1.26) | 0.055 |

** based on cluster‑robust (CR2) standard errors using small‑sample adjustments implemented in the clubSandwich package in R (69, 70)*

**Table S7.** Unconditional and detrended intraclass correlation coefficients for public stigma and perceived stigma, under progressive trimming of inverse probability of censoring weights

| **Measure** | **Unweighted*** | **IPC-weighted, untrimmed*** | **IPC-weighted, trimmed 1st/99th*** | **IPC-weighted, trimmed 5th/95th*** | **IPC-weighted, trimmed 10th/90th*** |
| --- | --- | --- | --- | --- | --- |
| Public stigma |  |  |  |  |  |
| ICC | 0.27 (0.25, 0.28) | 0.42 (0.39, 0.45) | 0.42 (0.39, 0.45) | 0.42 (0.39, 0.45) | 0.42 (0.39, 0.45) |
| ICC (detrended) | 0.35 (0.32, 0.37) | 0.49 (0.46, 0.52) | 0.49 (0.46, 0.52) | 0.49 (0.46, 0.52) | 0.49 (0.46, 0.52) |
| Perceived stigma |  |  |  |  |  |
| ICC | 0.00 (0.00, 0.00) | 0.11 (0.10, 0.13) | 0.11 (0.10, 0.13) | 0.11 (0.10, 0.13) | 0.11 (0.10, 0.12) |
| ICC (detrended) | 0.01 (0.00, 0.03) | 0.21 (0.19, 0.23) | 0.21 (0.19, 0.23) | 0.21 (0.19, 0.23) | 0.21 (0.19, 0.23) |

** 95% confidence intervals in brackets; based on nonparametric cluster bootstrap with 1,000 replications, resampling villages with replacement to account for clustering of participants within villages (68)*

**Figure S1.** Flow diagram depicting the open cohort design

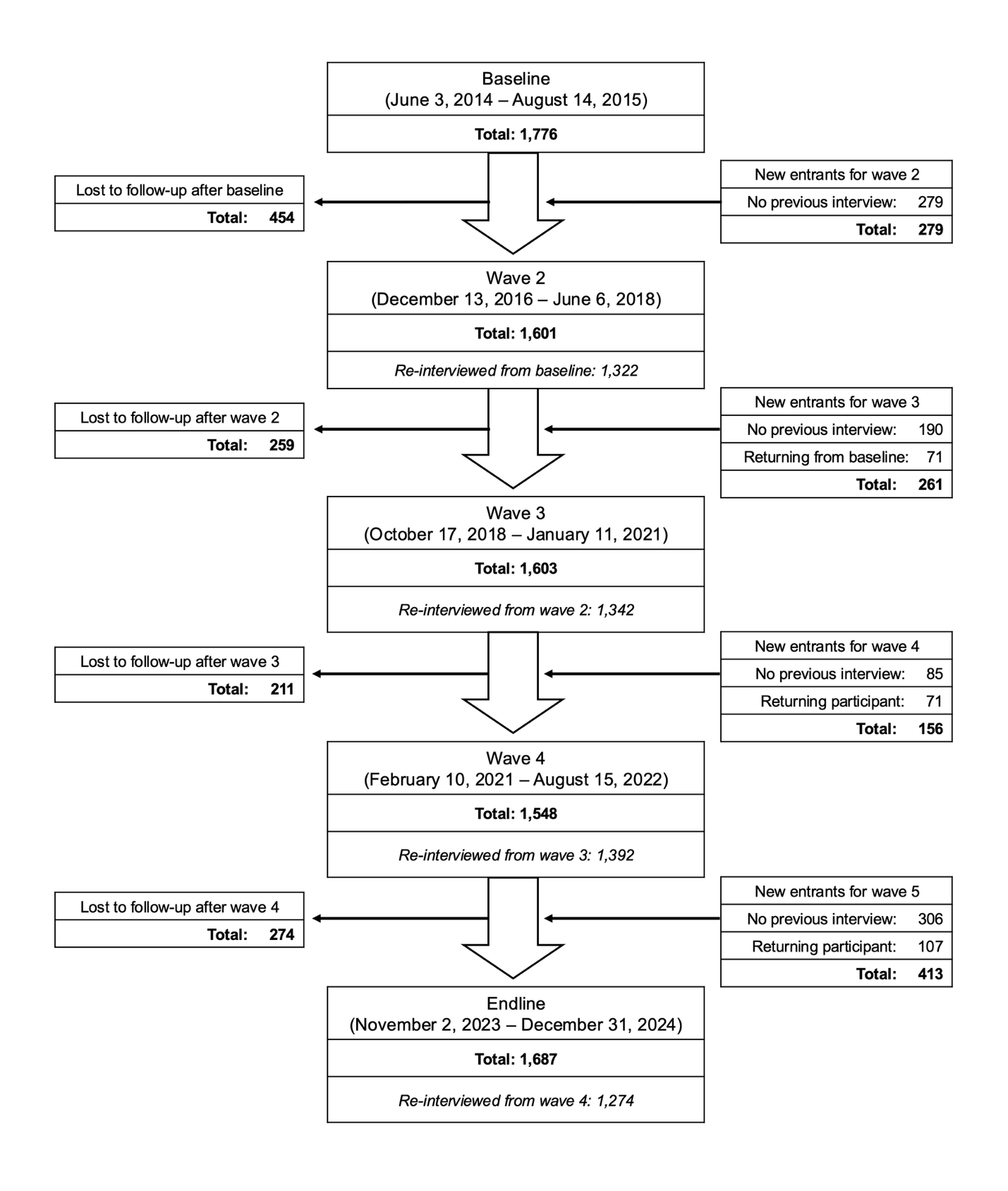

**Figure S2.** Distribution of HIV stigma scores, by survey wave

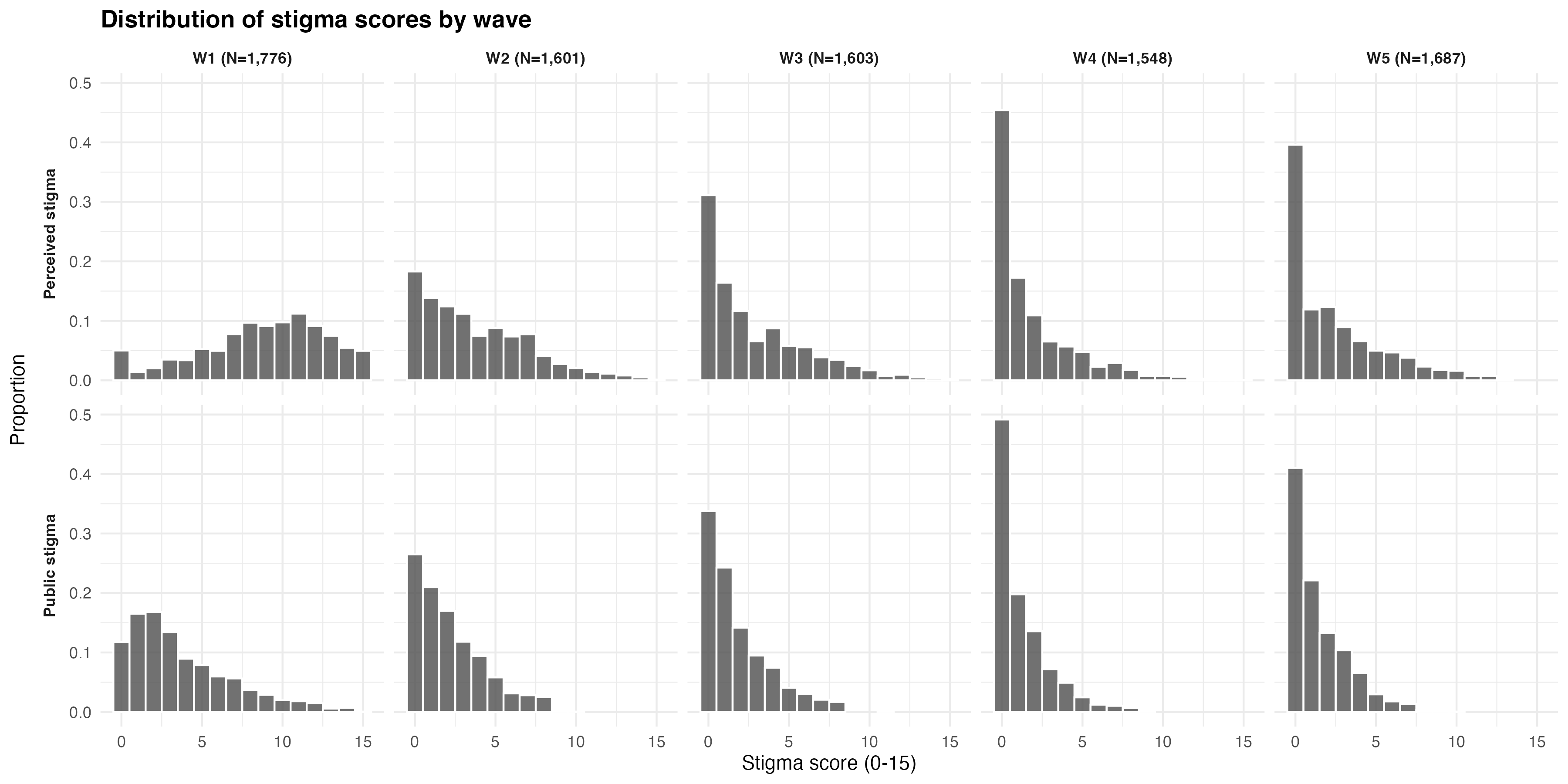

**Figure S3.** Trends in mean stigma and concentration, comparing estimates from the full serial cross sections vs. balanced panel only

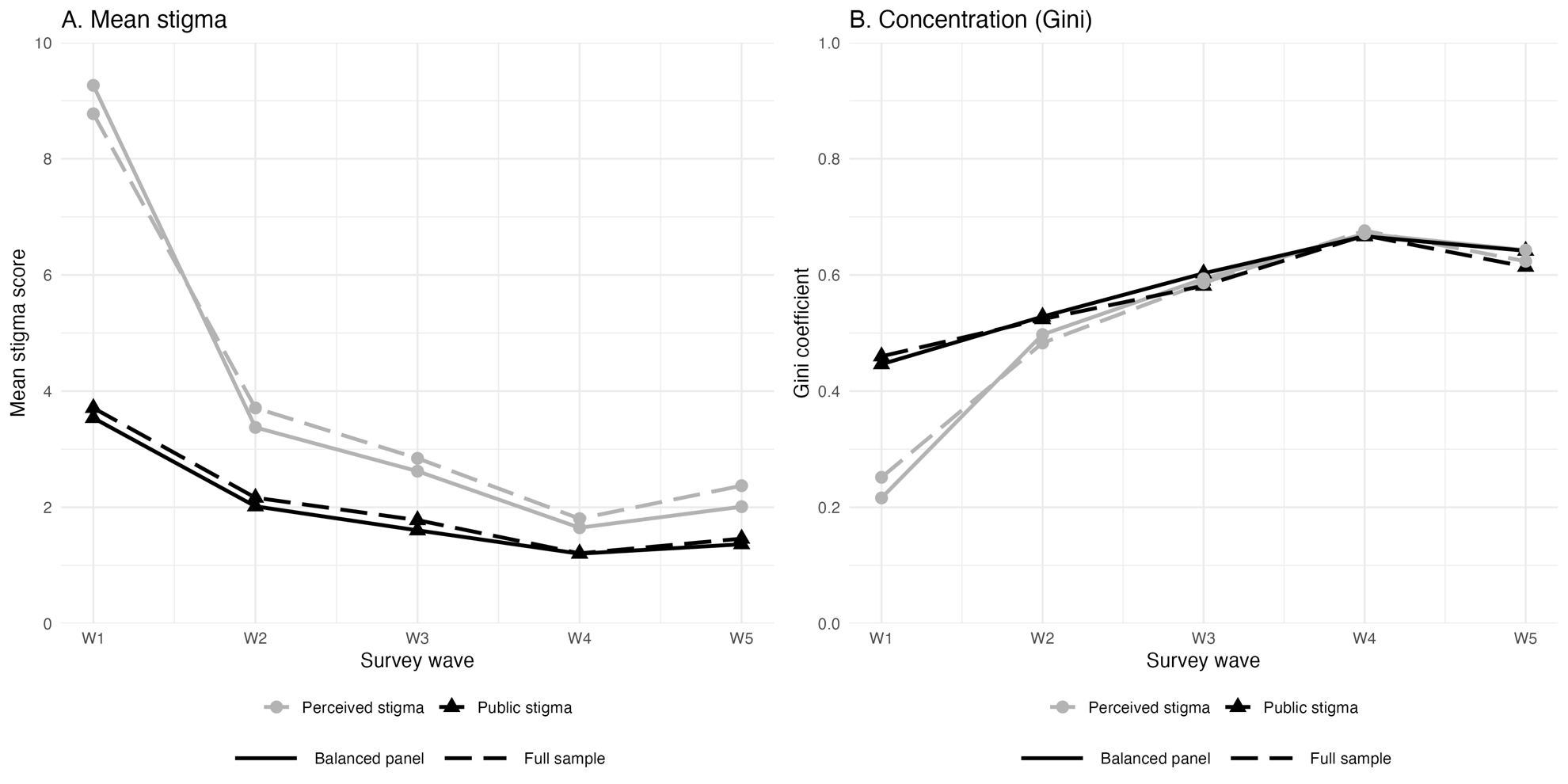

**Figure S4.** Rank-order stability across survey waves, after application of inverse probability of censoring weights

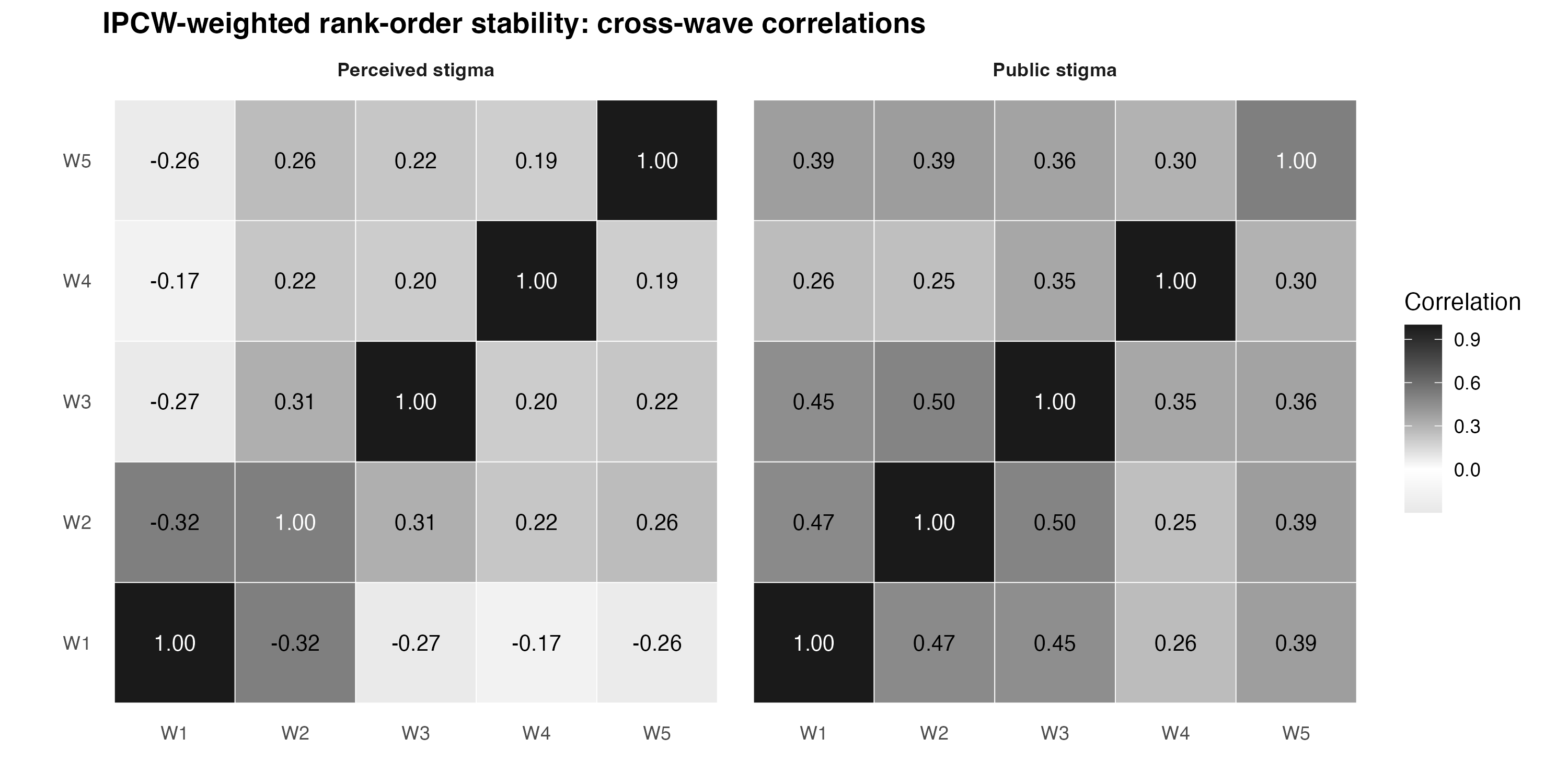

References

Andrews, F. M., & Withey, S. B. (1976). *Social indicators of well-being: Americans' perceptions of life quality*. New York: Plenum Press.

Bolton, P., & Ndogoni, L. (2001). Cross-cultural assessment of trauma-related mental illness (Phase II): a report of research conducted by World Vision Uganda and The Johns Hopkins University. Available at: <http://www.certi.org/publications/policy/ugandafinahreport.htm>. Last accessed April 23, 2011.

Borsari, B., & Carey, K. B. (2003). Descriptive and injunctive norms in college drinking: a meta-analytic integration. *J Stud Alcohol,* 64(3), 331–341.

Burt, R. S. (1984). Network items in the General Social Survey. *Soc Networks,* 6(4), 293–339.

Cahill, L., Gifford, A. J., Jones, B. A., & McDermott, D. T. (2025). The HIV Anxiety Scale (HAS): developing and validating a measure of Human Immunodeficiency Virus (HIV) anxiety. *AIDS Behav,* 29(7), 2258–2271.

Cole, S. R., & Hernán, M. A. (2008). Constructing inverse probability weights for marginal structural models. *Am J Epidemiol,* 168(6), 656–664.

Corrigan, P. W., & Penn, D. L. (1999). Lessons from social psychology on discrediting psychiatric stigma. *Am Psychol,* 54(9), 765–776.

Cowell, F. A., & Flachaire, E. (2007). Income distribution and inequality measurement: The problem of extreme values. *J Econometrics,* 141(2), 1044–1072.

de Walque, D. (2007). How does the impact of an HIV/AIDS information campaign vary with educational attainment? Evidence from rural Uganda. *J Dev Econ,* 84(2), 686–714.

Fatch, R., Bellows, B., Bagenda, F., Mulogo, E., Weiser, S., & Hahn, J. A. (2013). Alcohol consumption as a barrier to prior HIV testing in a population-based study in rural Uganda. *AIDS Behav,* 17(5), 1713–1723.

Gakidou, E. E., Murray, C. J., & Frenk, J. (2000). Defining and measuring health inequality: an approach based on the distribution of health expectancy. *Bull World Health Organ,* 78(1), 42–54.

Gurin, G., Veroff, J., & Feld, S. (1960). *Americans view their mental health: a nationwide interview survey*. New York: Basic Books, Inc.

Hernán, M. A., & Robins, J. M. (2006). Estimating causal effects from epidemiological data. *J Epidemiol Community Health,* 60(7), 578–586.

Kakuhikire, B., Satinsky, E. N., Baguma, C., Rasmussen, J. D., Perkins, J. M., Gumisiriza, P., et al. (2021). Correlates of attendance at community engagement meetings held in advance of bio-behavioral research studies: A longitudinal, sociocentric social network study in rural Uganda. *PLoS Med,* 18(7), e1003705.

Kipp, A. M., Audet, C. M., Earnshaw, V. A., Owens, J., McGowan, C. C., & Wallston, K. A. (2015). Re-validation of the Van Rie HIV/AIDS-related stigma scale for use with people living with HIV in the United States. *PLoS One,* 10(3), e0118836.

Kishor, S., & Johnson, K. (2004). *Profiling domestic violence: a multi-country study*. Calverton: ORC Macro.

Lapinski, M. K., & Rimal, R. N. (2005). An explication of social norms. *Comm Theory,* 15(2), 127–147.

Murphy, E. M., Greene, M. E., Mihailovic, A., & Olupot-Olupot, P. (2006). Was the "ABC" approach (abstinence, being faithful, using condoms) responsible for Uganda's decline in HIV? *PLoS Med,* 3(9), e379.

Mushavi, R. C., Burns, B. F. O., Kakuhikire, B., Owembabazi, M., Vořechovská, D., McDonough, A. Q., et al. (2020). "When you have no water, it means you have no peace": A mixed-methods, whole-population study of water insecurity and depression in rural Uganda. *Soc Sci Med,* 245, 112561.

NHLBI Obesity Education Initiative Expert Panel on the Identification, E., and Treatment of Obesity in Adults, (1998). Clinical guidelines on the identification, evaluation, and treatment of overweight and obesity in adults—the evidence report. National Institutes of Health. *Obes Res,* 6, 51S–209S.

Nyblade, L., MacQuarrie, K., Phillip, F., Kwesigabo, G., Mbwambo, J., Ndega, J., et al. (2005). *Measuring HIV stigma: results of a field test in Tanzania*. Washington, D.C.: U.S. Agency for International Development.

Pradhan, M., Sahn, D. E., & Younger, S. D. (2003). Decomposing world health inequality. *J Health Econ,* 22(2), 271–293.

Pronyk, P. M., Hargreaves, J. R., Kim, J. C., Morison, L. A., Phetla, G., Watts, C., et al. (2006). Effect of a structural intervention for the prevention of intimate-partner violence and HIV in rural South Africa: a cluster randomised trial. *Lancet,* 368(9551), 1973–1983.

Robins, J. M., Hernán, M. A., & Brumback, B. (2000). Marginal structural models and causal inference in epidemiology. *Epidemiology,* 11(5), 550–560.

Tsai, A. C., Kakuhikire, B., Perkins, J. M., Vořechovská, D., McDonough, A. Q., Ogburn, E. L., et al. (2017). Measuring personal beliefs and perceived norms about intimate partner violence: Population-based survey experiment in rural Uganda. *PLoS Med,* 14(5), e1002303.

U.S. National Center for Health Statistics (1996). *NHANES III Anthropometric Procedures Video. U.S. Government Printing Office Stock Number 017-022-01335-5. Available from:* [*https://www.ncbi.nlm.nih.gov/books/NBK2004/box/A236/*](https://www.ncbi.nlm.nih.gov/books/NBK2004/box/A236/). Washington, D.C.: U.S. Government Printing Office.

VanderWeele, T. J., & Ding, P. (2017). Sensitivity analysis in observational research: introducing the E-value. *Ann Intern Med,* 167(4), 268–274.

Wagner, A. C., Hart, T. A., McShane, K. E., Margolese, S., & Girard, T. A. (2014). Health care provider attitudes and beliefs about people living with HIV: Initial validation of the Health Care Provider HIV/AIDS Stigma Scale (HPASS). *AIDS Behav,* 18(12), 2397–2408.
